# Artificial Intelligence to Assist in the Screening Fetal Anomaly Ultrasound Scan (PROMETHEUS): A Randomised Controlled Trial

**DOI:** 10.1101/2024.05.23.24307329

**Authors:** Thomas G Day, Jacqueline Matthew, Samuel F Budd, Alfonso Farruggia, Lorenzo Venturini, Robert Wright, Babak Jamshidi, Meekai To, Huazen Ling, Jonathon Lai, Min Yi Tan, Matthew Brown, Gavin Guy, Davide Casagrandi, Anastasija Arechvo, Argyro Syngelaki, David Lloyd, Vita Zidere, Trisha Vigneswaran, Owen Miller, Ranjit Akolekar, Surabhi Nanda, Kypros Nicolaides, Bernhard Kainz, John M Simpson, Jo V Hajnal, Reza Razavi

## Abstract

**Background:** Artificial intelligence (AI) has shown potential in improving the performance of screening fetal anomaly ultrasound scans. We aimed to assess the effect of AI on fetal ultrasound scanning, in terms of diagnostic performance, biometry, scan duration, and sonographer cognitive load.

**Methods:** This was a randomised, single centre, open label trial in a large teaching hospital. Pregnant participants with fetal congenital heart disease (CHD) and with healthy fetuses were recruited and scanned with both methods. Screening sonographers were recruited from regional hospitals and were randomised to scan with the AI tool or in the standard fashion, blinded to the fetal CHD status. For the AI-assisted scans, the AI models identified and saved 13 standard image planes, and measured four biometrics.

**Findings:** 78 pregnant participants (26 with fetal CHD) and 58 sonographers were recruited. The sensitivity and specificity of the AI-assisted scan in detecting fetal malformation was 88.9% and 98.0% respectively, with the standard scan achieving 81.5% and 92.2% (not significant). AI-assisted scans were significantly shorter than standard scans (median 11.4 min vs 19.7 min, p <0.001). Sonographer cognitive load was significantly lower in the AI-assisted group (median NASA TLX score 35.2 vs 46.5, p <0.001). For all biometrics, the AI repeatability and reproducibility was superior to manual measurements.

**Interpretation:** AI assistance in the routine fetal anomaly ultrasound scan results in a significant time saving, along with a reduction in sonographer cognitive load, without a reduction in diagnostic performance.

**Funding:** The study was funded by an NIHR doctoral fellowship (NIHR301448) and was supported by grants from the Wellcome Trust (IEH Award, 102431), by core funding from the Wellcome Trust/EPSRC Centre for Medical Engineering (WT203148/Z/16/Z), and the London AI Centre for Value Based Healthcare via funding from the Office for Life Sciences.

**Research in context:** *Evidence before this study:* We undertook a systematic review exploring evidence for the use of artificial intelligence (AI) to assist in the performance of fetal anomaly ultrasound scans by automating anatomical standard plane detection, and / or automating fetal biometric measurements. We searched PubMed, the Cochrane Library, and clinicaltrials.gov databases using the following search terms: ((artificial intelligence) OR (AI) OR (machine learning) OR (neural network*) OR (deep learning)) AND ((fetal) OR (foetal) OR (fetus) OR (foetus) OR (obstetric) OR (antenatal) OR (prenatal) OR (pregnan*)) AND (ultrasound) AND ((biometr*) OR (measurement) OR (growth) OR (size) OR (plane) OR (view) OR (femur) OR (head) OR (biparietal) OR (abdom*)). We limited the results to the 10-year period September 2013 – September 2023. 770 papers were identified, of which 55, 39, and 33 were deemed relevant after title, abstract, and text screening respectively. Of the 33 papers, 14 focused on biometric measurements, 14 on plane detection, and 5 included both. Only one paper tested the AI models prospectively with real-time feedback to the sonographer, but this study did not include randomisation or fetal pathology. No randomised clinical trials comparing AI-assisted ultrasound scans with standard scans have been performed previously.

*Added value of this study:* To our knowledge, this is the first randomised controlled trial investigating the use of AI in fetal ultrasound screening. In this trial we assess the use of AI-assistance to automatically undertake some aspects of the scan (automatic anatomical standard plane detection and saving, and measurement of biometric parameters), and measure the effect on overall diagnostic performance, as well as scan duration, sonographer cognitive load, image quality, and repeatability and reproducibility of biometric measurements. AI assistance resulted in a significantly lower scan duration and sonographer cognitive load, whilst maintaining the quality of the scan in terms of diagnostic performance and biometric measurements.

*Implications of all the available evidence:* The results from this trial are encouraging and suggest that AI assistance may offer real clinical benefit to sonographers undertaking fetal ultrasound screening. The reduced scan duration means that sonographers may have more time to focus on other aspects of the scan, such as communication with parents. The automatically measured biometrics were both more repeatable and reproducible compared to manual measurements, which may improve the accuracy with which fetal growth and health can be assessed. Further studies combining this work with AI models that can directly detect fetal structural malformations will be important, to improve the overall antenatal detection of fetal anomalies.

## Background

Congenital malformations are the most common causes of infant mortality in high-income countries such as the UK and USA, and are becoming increasingly important worldwide as other causes of child death become less common.^1^ Antenatal diagnosis is desirable as it has been shown to reduce postnatal mortality and morbidity for some lesions, may lead to therapeutic intervention in selected cases and allows the parents to make an informed decision about whether they wish the pregnancy to continue.^2,3^ The mainstay of antenatal diagnosis is the fetal anomaly screening ultrasound scan. In the UK, the Fetal Anomaly Screening Programme (FASP) stipulates an offer of this scan between 18^+0^ and 20^+6^ weeks gestation, with the aim of detecting 11 specific fetal conditions.^4^

Despite very high rates of uptake for these scans, universal detection of major fetal malformations has not been achieved. For example, in the UK only 50.4% of infants undergoing surgery for congenital heart disease (CHD) have received an antenatal diagnosis, and there is also wide regional variation across the country.^5^ Artificial intelligence has been postulated as a means to improve the performance of many medical tasks, including the fetal anomaly ultrasound scan.^6^ Previous studies have described the development of AI models to automate aspects of the scan such as plane detection and fetal biometry ^7–11^, including a pilot study by our group examining prospective real-time use with normal fetuses.^6^

However, in many branches of medicine, including obstetrics, the recent explosion of interest in AI has not been accompanied by high-quality prospective clinical trials.^12^ Despite a large literature describing good model performance when tested on retrospective ultrasound data, no prospective randomised trial examining the real-world effect of AI on the fetal anomaly scan, including abnormal fetuses, has yet been published.

We have created a clinical tool that combines AI models to automate plane detection and image saving, and measurement of fetal biometric parameters, in real-time, with live feedback to the sonographer. This tool fundamentally alters the way in which the scan is performed, as sonographers no longer need to pause, save images, or measure during the scan, resulting in fewer interruptions and a more streamlined workflow. By automating some aspects of the ultrasound examination, we hypothesised that the sonographers would be able to improve their detection of fetal malformation.

Our aim was to undertake a randomised controlled trial examining the effects of this tool in a population including fetuses with known major structural malformations, involving sonographers from a variety of professional backgrounds. We selected CHD as the focus of the study as it is the group of congenital malformations that is most common, most commonly missed, and has the highest infant mortality.^13,14^ The outcome measures for the trial were sensitivity and specificity for CHD detection, the duration of the scan, and the cognitive load of the sonographers, as well as the quality of saved images, and repeatability and reproducibility of automated fetal measurements. We have employed a novel trial design for low prevalence disease, with a study population enriched with fetuses affected by CHD, allowing assessment of diagnostic performance in a reasonable sample size. Variation between study participants (both pregnant participants and sonographers) was controlled for, as all pregnant participants underwent both an AI-assisted scan and a standard manual scan as comparison, with randomisation of the sonographers performing each type of scan.

## Methods

### Study design and participants

The PROMETHEUS trial (Prospective tRial of Machine lEarning To Help fEtal Ultrasound Scanning) was a single-centre randomised controlled open-label trial of AI-assisted vs. standard unassisted fetal anomaly ultrasound scans. The study was designed so that on a given study day three pregnant participants (two with a fetus with a normal heart, one with a fetus with CHD) were invited to attend the study site, along with two sonographers, who were randomised to perform scans either with or without AI assistance. Each pregnant participant was scanned twice sequentially, once using each method. The scans were research investigations, and not intended to perform a clinical purpose, and performed in addition to standard clinical investigations, thus standard care was not affected by participation in the trial. The trial was conducted in the Clinical Research Facility of a large urban teaching hospital in the UK. The study protocol was prospectively registered with ISRCTN, number 65824874.

Pregnant participants were recruited from a tertiary centre of fetal cardiology, either following a diagnosis of fetal congenital heart disease (“affected group”), or in whom the fetus had been confirmed to have a normal heart structure after detailed fetal echocardiography (“unaffected group”). The unaffected group had been offered detailed fetal cardiac screening because of a risk factor for CHD, such as family history, maternal diabetes, or drug exposure. Inclusion criteria were either diagnosis of fetal CHD between 12^+0^ and 27^+6^ weeks gestation (for the affected group) or confirmation of normal fetal cardiac anatomy between 18^+0^ and 27^+6^ weeks gestation (for the unaffected group), with at least one week between CHD diagnosis and recruitment, if present. Exclusion criteria for pregnant participants were: any plan for termination of pregnancy; any known fetal extracardiac structural abnormality at the time of recruitment; any known fetal genetic abnormality; multiple pregnancy; refusal of consent; insufficient English language skills to provide informed consent; or age under 18. Potential participants were contacted by telephone, after approval from the clinical specialist nursing team caring for the patient. The research anomaly scans were performed between 18^=0^ - 27^+6^ weeks’ gestation.

Sonography professionals were recruited from units within South East England, via email invitation to the sonographer lead at each department, with a request for them to cascade to their staff. Advertisements were also placed in electronic newsletters of professional groups (British Medical Ultrasound Society and The Society of Radiographers). The inclusion criterion was the regular independent performance of fetal anomaly screening ultrasound scans as part of their clinical work. Exclusion criteria were any previous involvement in our research projects, or refusal of consent. Written consent was obtained from both pregnant participants and sonographers. They could be from any professional background (e.g. radiography, midwifery, nursing, or medical), and all were termed ‘sonographers’ for the purposes of this study.

Ethics approval was granted by the London Dulwich Research Ethics Committee, reference 22/LO/0163.

### Randomisation and masking

Randomisation was performed at a sonographer level on the day of the study session. The pair of sonographers attending for that study session were randomised such that one performed the scan with AI assistance, and the other performed a standard manual scan. An online tool was used for the randomisation (randomizer.org). The sonographers were blinded to the clinical status of the pregnant participants (i.e. healthy or CHD), and the fact that CHD was the focus of the study.

### Procedures

The AI models used in this study performed two tasks: 1) the detection and labelling of standard image planes from a stream of ultrasound video, and 2) the automatic fetal biometry. The models were developed using a prospectively acquired dataset of 7,309 complete videos of routine anomaly ultrasound scans performed in a single institution. The training and testing of the AI models are described in supplementary information 1, and described more fully in Venturini *et al* and Baumgartner *et al.*^10,18^ The standard planes and biometrics used in the study are shown in Table 1, based on the UK Fetal Anomaly Screening Programme (FASP).^7^

**Table 1:**
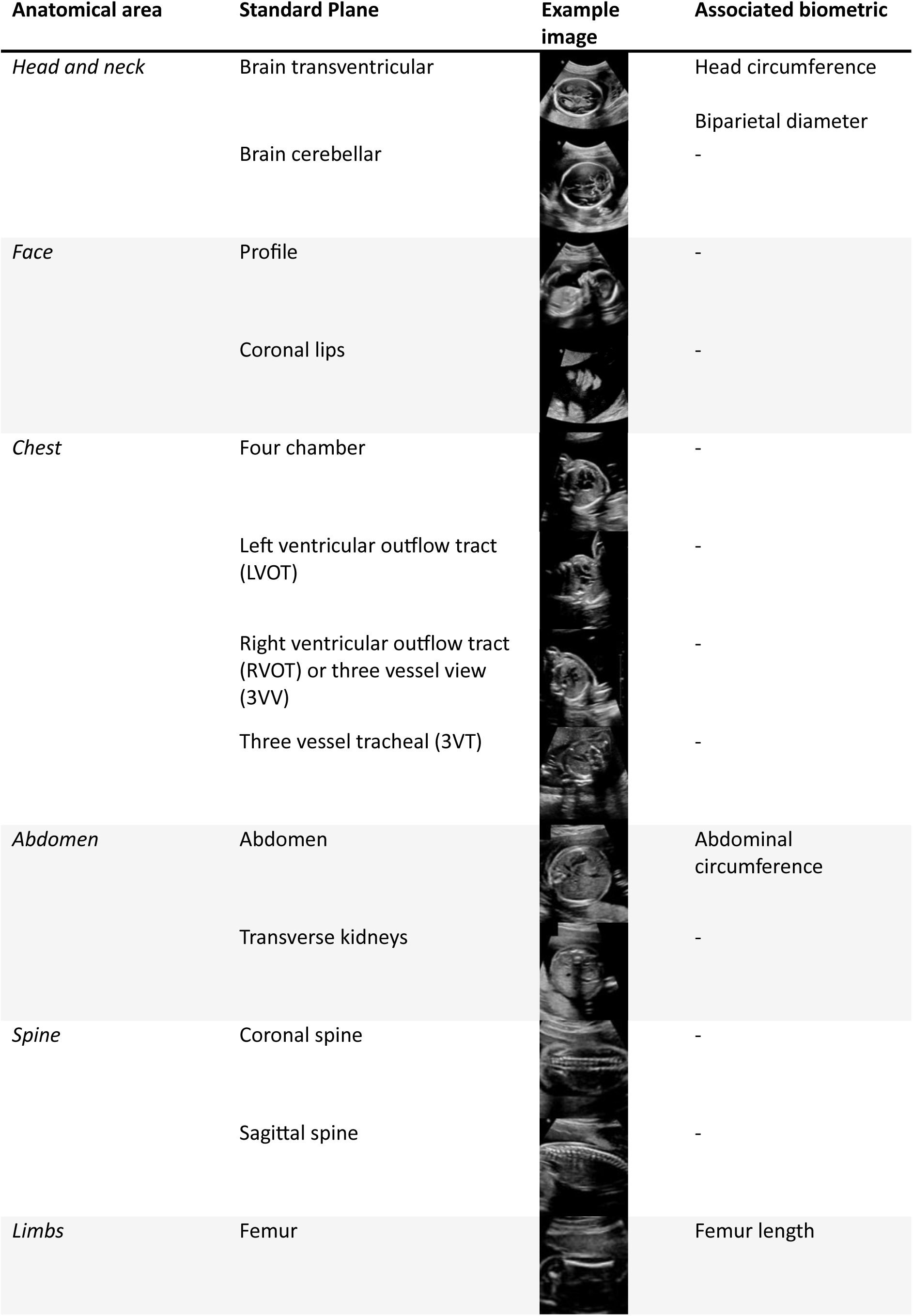
standard fetal ultrasound image planes and associated biometric measurements used in the study.

A GE Voluson Expert 22 ultrasound machine was used for the study. Our clinical AI tool consisted of a computer (Boxer-8641AI, Aaeon Technology Inc., Taipei, Taiwan) mounted on the ultrasound machine, receiving as an input the stream of ultrasound video via a high-definition multimedia interface (HDMI) connection. The individual images within the video stream were analysed in real time by our AI models as described in supplementary information 1 and 2, with outputs immediately displayed to the sonographer via a tablet (iPad Air 5^th^ Generation, Apple Inc. Cupertino, USA). The tablet was connected to the computer via a Wi-Fi connection. A schematic diagram and photograph of the technical setup is shown in Figure 1.

**Figure 1:**
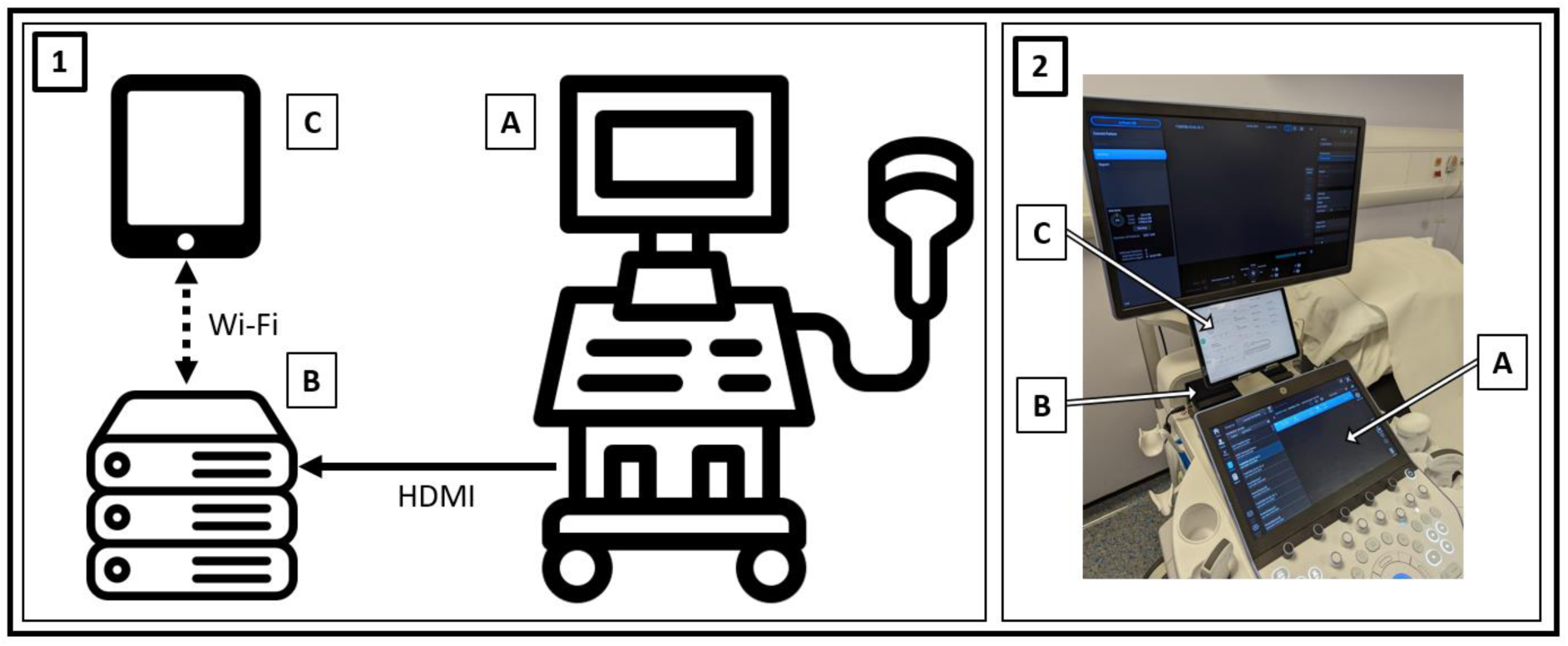
technical integration of artificial intelligence assistance tool. 1: schematic diagram of study setup. 2: photograph of study setup. A: standard ultrasound machine; B: computer mounted on ultrasound machine, receiving video stream of ultrasound scan via HDMI cable; C: tablet displaying outputs of AI models; HDMI: high-definition multimedia interface; solid black arrow: physical cable connection; dashed black arrow: connection via internet.

Sonographers underwent a 15 minute 1:1 training session on the day of the study with an investigator, with a written guide and video. They were asked to follow a study-specific scan protocol, shown in supplementary information 3. Sonographers performing the manual scan were asked to save a single image for each of the 13 standard planes. They were asked to measure each of the four biometric parameters three times and select the best measurement for their report (as per published guidance).^19^ For the sonographers performing the AI-assisted scan, saving of image planes and measurement of biometrics was performed automatically by the AI tool. Feedback on these processes was shown to the AI-assisted sonographer via the tablet. During the scan, the current best estimate for each biometric was displayed in real time to the sonographer on a scale indicating the normal values for the given gestation, along with a calculated error bound. Similarly, for each standard plane, a labelled progress bar indicated how many images had been saved. For the AI-assisted scan, after completion the sonographers were shown a candidate image for each of the standard planes on the tablet. They could either accept this image or choose from a further eight images for each plane to be selected as their final best image (the “image review” stage).

After each scan, the sonographers completed a written report on a standard laptop using a web-based interface. They were asked to choose a final outcome from a choice of a) standard follow up (i.e. usual antenatal care), b) a repeat scan in a screening department (e.g. because of poor views of a certain anatomical area because of fetal position), or c) referral to specialist services (e.g. because CHD had been identified). They also completed survey instruments to measure cognitive load using both the NASA-TLX scale and Paas scale.^20,21^ The NASA-TLX scale was used unweighted as previously described, and is a multidimensional instrument with six subscales (mental, physical, and temporal demands, and frustration, effort, and performance), designed to capture different aspects of cognitive load, each recorded using a visual analogue scale and then summed.^22^ The Paas scale is a 9-point Likert scale response to the question “please rate your mental effort required to perform the scan”.

After the end of the trial, the quality of each saved image was assessed by experts in fetal medicine, blinded to the method used to acquire each image. A quality scoring scheme was agreed by consensus with the experts prior to this, with further details found in supplementary information 4. This resulted in two metrics for each image: a binary outcome indicating if the image was deemed ‘clinically acceptable’ or not, and a further continuous outcome indicating overall image quality, normalised to a scale from 0-1. Further AI models that could automatically discriminate the quality of saved images became available after the trial had commenced. To examine the effectiveness of these, all saved images from the AI scans were passed through these models, with the top nine images displayed to a research sonographer (as in the live trial). The best quality image from these nine was selected by an experienced research sonographer, and scored for quality in the same way as the images selected initially during the study sessions. This resulted in a single image being chosen and graded per plane per participant for each of the three methods (manual acquisition, AI acquisition, and AI acquisition with retrospective use of quality models).

### Outcomes

The primary outcome measures were the sensitivity and specificity of the two methods in detecting CHD. A scan was defined as positive for CHD if at least one of the cardiac views was described as abnormal or not seen in the written report, and the final outcome of the scan was a referral to specialist services. All other scans were defined as negative for CHD. This was compared to the ground truth to classify all scans as true positive, false positive, true negative, or false negative for fetal CHD. If an unaffected fetus was unexpectedly identified as being suspected of having CHD an urgent repeat specialist fetal echocardiogram was performed on the same day to define whether this was a true or false positive finding. Secondary outcome measures were the time taken to complete the scan and report, the cognitive load of the sonographers, the quality of saved images, and the repeatability and reproducibility of fetal biometrics. All pregnant participants were followed up after delivery to confirm that the antenatal diagnoses were correct.

### Statistical analysis

The sample size gave an 80% power to demonstrate non-inferiority of CHD detection sensitivity with a target of 80% and delta of 25%. Groups were compared using Wilcoxon signed rank test for paired continuous data, Mann-Whitney U test for unpaired continuous data, and McNemar’s test for paired proportions. 95% confidence intervals for proportions were calculated using the exact Clopper-Pearson method. Statistical analysis was performed using SPSS version 29.0.0.0 (IBM Corporation, Armonk, USA). A *p* value of less than 0.05 was considered significant.

Manual and AI biometrics were compared using Bland-Altman plots. The three measurements per biometric recorded during the manual scan were used to calculate the repeatability of the manual method (since the mean of distance between the maximum and minimum of n observations for a uniform distribution over interval (a b) is equal to ((n-1)/(n+1))*(b-a), by using the coefficient 2/3 a balance was made between these three-measurement criteria, and the other comparison which had only two measurements). The chosen ‘best’ manual measurement was compared with the final estimate from the AI-assisted scan to compare reproducibility between the two methods. The mean difference between the two methods was also subtracted from the AI measurement, so that the random error could be visualised (i.e. removing any systematic bias). Manual human reproducibility (interobserver variability) was not measured in this trial design, but this has been published previously for three of the four biometrics.^23^ Finally, the video recorded during the manual scan was analysed by the AI model retrospectively to obtain a second AI biometric measurement on the same patient on a sequential scan, so that AI-AI repeatability could be assessed. Figure 2 shows a diagram of how these measurements were compared.

**Figure 2:**
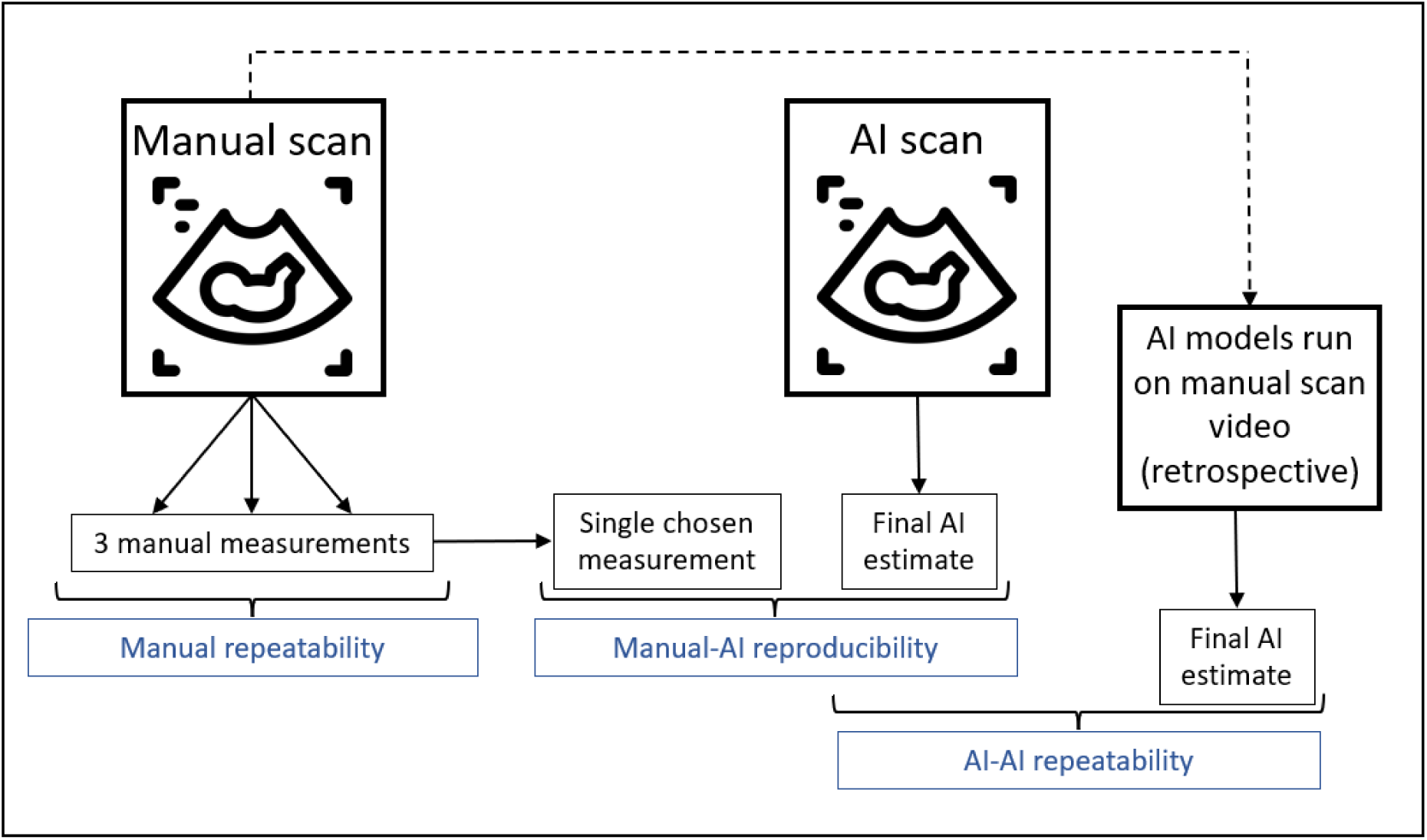
schematic diagram describing comparison of the biometric measurements.

## Results

Figure 3 shows recruitment figures for sonographers, with 58 recruited over the period 05/05/2022 – 17/07/23. 29 sonographers were randomised into each scanning method. Baseline characteristics after randomisation are shown in Table 2. Figure 4 shows recruitment figures for pregnant participants, with 78 recruited over the period 17/11/22 – 01/08/23 and included in the final analyses. Baseline characteristics are shown in Table 3, with details of the CHD lesions in cases in Table 4. The study sessions ran from 15/11/22 – 08/08/23.

**Figure 3:**
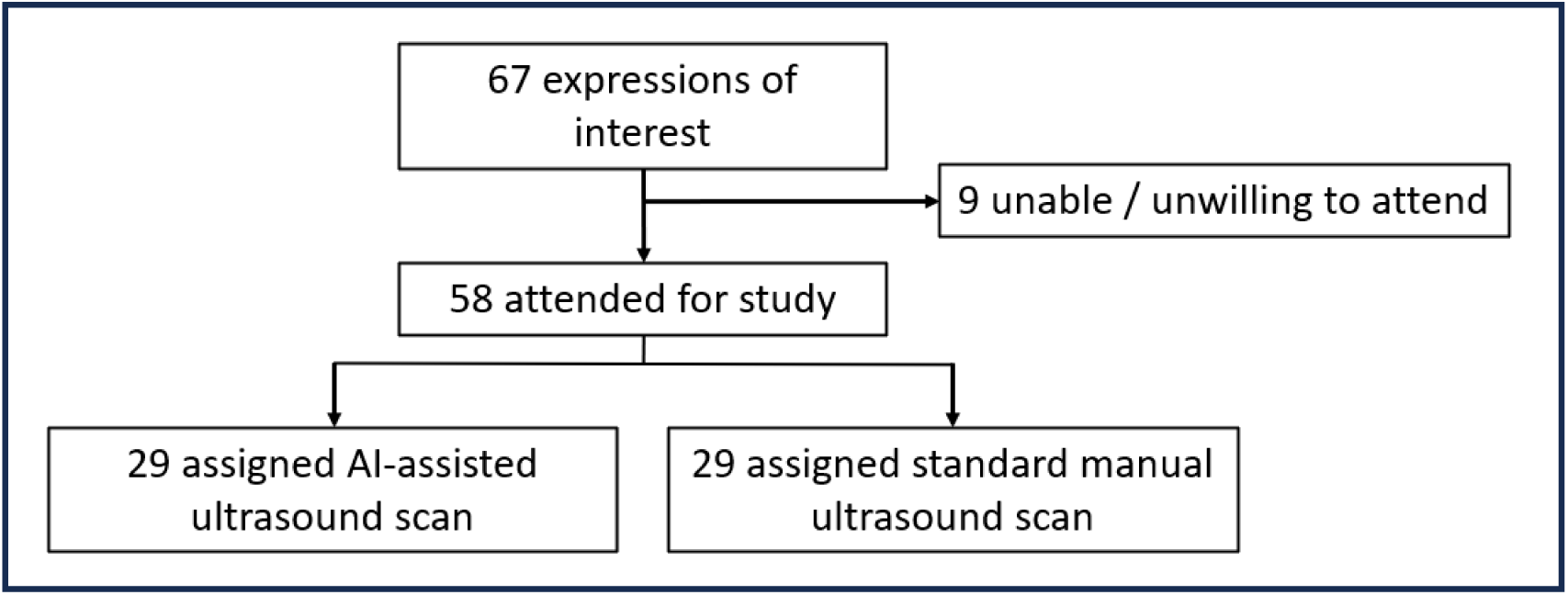
recruitment flowchart for sonographer participants.

**Figure 4:**
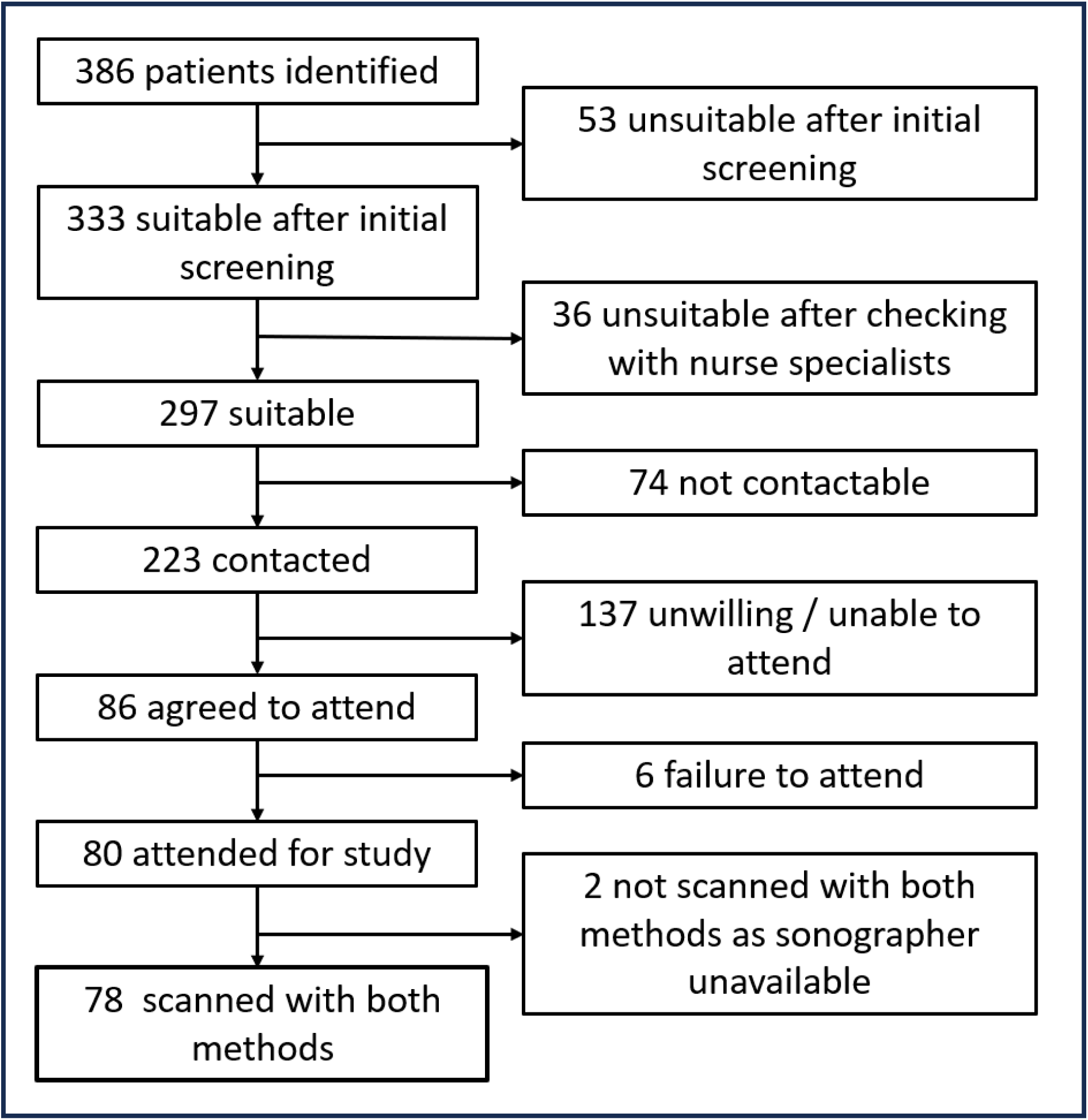
recruitment flowchart for pregnant participants.

**Table 2:** baseline characteristics of sonographer participants after randomisation. Data are n (%) or median (IQR).

|  | AI-assisted scan (n=29) | Manual scan (n=29) |
| --- | --- | --- |
| **Experience in fetal ultrasound (years)** | 4 (3-10) | 6 (2-12) |
| **Professional background** |  |  |
| Radiographer | 13 (44.8%) | 13 (44.8%) |
| Nurse / midwife | 3 (10.3%) | 0 |
| Doctor | 13 (44.8%) | 16 (55.2%) |

**Table 3:** baseline characteristics of pregnant participants. Data are n (%) or mean (SD).

|  | CHD affected group | CHD unaffected group |
| --- | --- | --- |
| **Maternal age (years)** | 32.1 (5.4) | 31.2 (6.0) |
| **Gestational age (weeks)** | 25.3 (1.9) | 24.9 (1.6) |
| **Body mass index (kg/m<sup>2</sup>)** | 27.9 (5.4) | 27.6 (4.6) |
| **Ethnicity** |  |  |
| White | 20 (76.9%) | 51 (94.4%) |
| Black or mixed black | 3 (11.5%) | 1 (1.9%) |
| Asian or mixed Asian | 1 (3.8%) | 2 (3.7%) |
| Any other | 2 (7.7%) | 0 |
| **Reason for referral to fetal cardiology** |  |  |
| Suspected CHD | 24 (92.3%) | 3 (5.6%) |
| Family history of CHD | 0 | 34 (63.0%) |
| Maternal diabetes | 0 | 5 (9.3%) |
| Teratogenic medications | 1 (3.8%) | 3 (5.6%) |
| Raised nuchal translucency | 1 (3.8%) | 6 (11.1%) |
| Other | 0 | 3 (5.6%) |

**Table 4:**
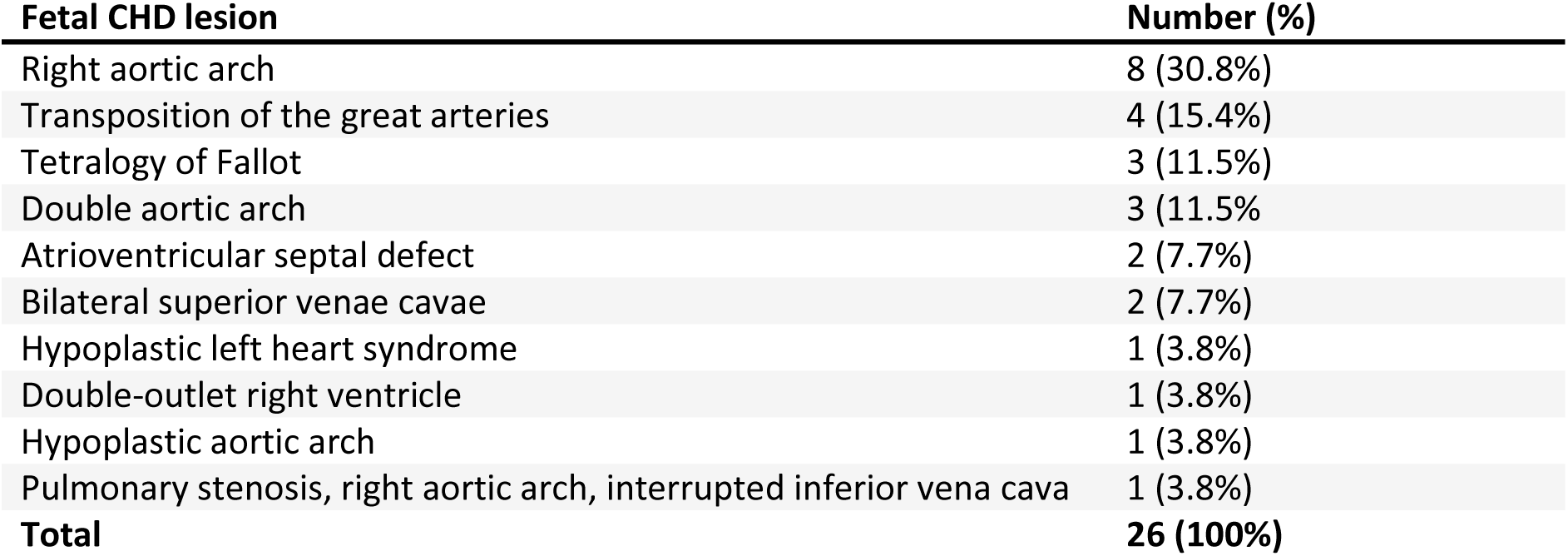
details of fetal CHD lesions in cases.

| Fetal CHD lesion | Number (%) |
| --- | --- |
| Right aortic arch | 8 (30.8%) |
| Transposition of the great arteries | 4 (15.4%) |
| Tetralogy of Fallot | 3 (11.5%) |
| Double aortic arch | 3 (11.5%) |
| Atrioventricular septal defect | 2 (7.7%) |
| Bilateral superior venae cavae | 2 (7.7%) |
| Hypoplastic left heart syndrome | 1 (3.8%) |
| Double-outlet right ventricle | 1 (3.8%) |
| Hypoplastic aortic arch | 1 (3.8%) |
| Pulmonary stenosis, right aortic arch, interrupted inferior vena cava | 1 (3.8%) |
| **Total** | **26 (100%)** |

**Table 5:** diagnostic performance of the two methods in detecting fetal congenital heart disease (affected group n=26, unaffected group n=52). * McNemar’s test for paired proportions.

|  | AI-assisted scan | Manual scan | P value* |
| --- | --- | --- | --- |
| True positive (n) | 21 | 20 | - |
| False positive (n) | 0 | 4 | - |
| True negative (n) | 52 | 48 | - |
| False negative (n) | 5 | 6 | - |
| Sensitivity (95% CI) | 80.8% (60.6-93.4%) | 76.9% (56.4-91.0%) | 0.705 |
| Specificity (95% CI) | 100% (93.2-100%) | 92.3% (81.5-97.9%) | **0.046** |

Although 156 paired scans were performed, data were not available from every scan for every outcome measure, because of prototype software or hardware failures during the study procedures. This is described in more detail in Supplementary Information 5.

The primary outcome measure for the trial was the diagnostic performance of the scan in detecting fetal CHD. There was no significant difference between the two methods in terms of sensitivity (AI-assisted scan 80.8%, manual scan 76.9%, p = 0.705), but the AI-assisted scan was significantly more specific for CHD than the manual scan (100% and 92.3% respectively, p = 0.046).

Postnatal outcome was available for 73 out of 78 fetuses (97.3%), with five not contactable after birth (all in the unaffected group). All fetuses in the CHD group had a postnatal diagnosis that was concordant with their antenatal cardiac diagnosis. Six fetuses in the group unaffected by CHD had a postnatal echocardiogram due to either a heart murmur on routine postnatal examination, or family history of CHD. Three of these found minor abnormalities that would not be considered detectable on routine antenatal screening (one small secundum atrial septal defect, one very mild pulmonary valve stenosis not requiring treatment, and one subtle hypertrophy of the left ventricle not requiring treatment), so they remained in the unaffected group for the purposes of analysis. The other three were entirely normal.

Although known fetal extracardiac abnormalities at time of recruitment was an exclusion criterion, two pregnant participants were included with extracardiac abnormalities as they were identified after recruitment but prior to the study session (one unilateral hydronephrosis, treated with conservatively after birth, and one talipes, treated surgically after birth), one of these was also affected by CHD. Both were identified by both scanning methods. In addition, there were four suspected abnormalities identified during the study scans that were found to be not present on subsequent expert review and after birth (two suspected talipes, one suspected echogenic bowel, and one suspected cleft lip), all suspected during the AI-assisted scan only. Three of these four had coexisting CHD. Table 6 shows an alternative analysis in which all fetal structural malformations (CHD plus extracardiac anomalies) are considered as the affected group. When analysed in this way there were no significant differences in sensitivity or specificity between the two scanning methods.

**Table 6:** diagnostic performance of the two methods in detecting all fetal structural malformations (affected group n=27, unaffected group n=51). * McNemar’s test for paired proportions.

|  | **AI-assisted scan** | **Manual scan** | **P value\*** |
| --- | --- | --- | --- |
| True positive (n) | 24 | 22 | - |
| False positive (n) | 1 | 4 | - |
| True negative (n) | 50 | 47 | - |
| False negative (n) | 3 | 5 | - |
| Sensitivity (95% CI) | 88.9% (70.8-97.6%) | 81.5% (61.9-93.7%) | 0.480 |
| Specificity (95% CI) | 98.0% (89.6-100%) | 92.2% (81.1-97.8%) | 0.180 |

Results for scan and reporting duration are shown in Table 7 and Figure 5. The tablet-based image review stage (unique to the AI-assisted scan) was included in reporting time. The median scan duration was significantly shorter for the AI-assisted scan, saving on average 8.3 minutes (equivalent to 42% of the median manual scan time (Table 7). There was no significant difference in reporting time between the two groups. Figure 5: duration of scanning and reporting by both methodsFigure 5 shows the distribution of durations for both scanning and reporting.

**Figure 5:**
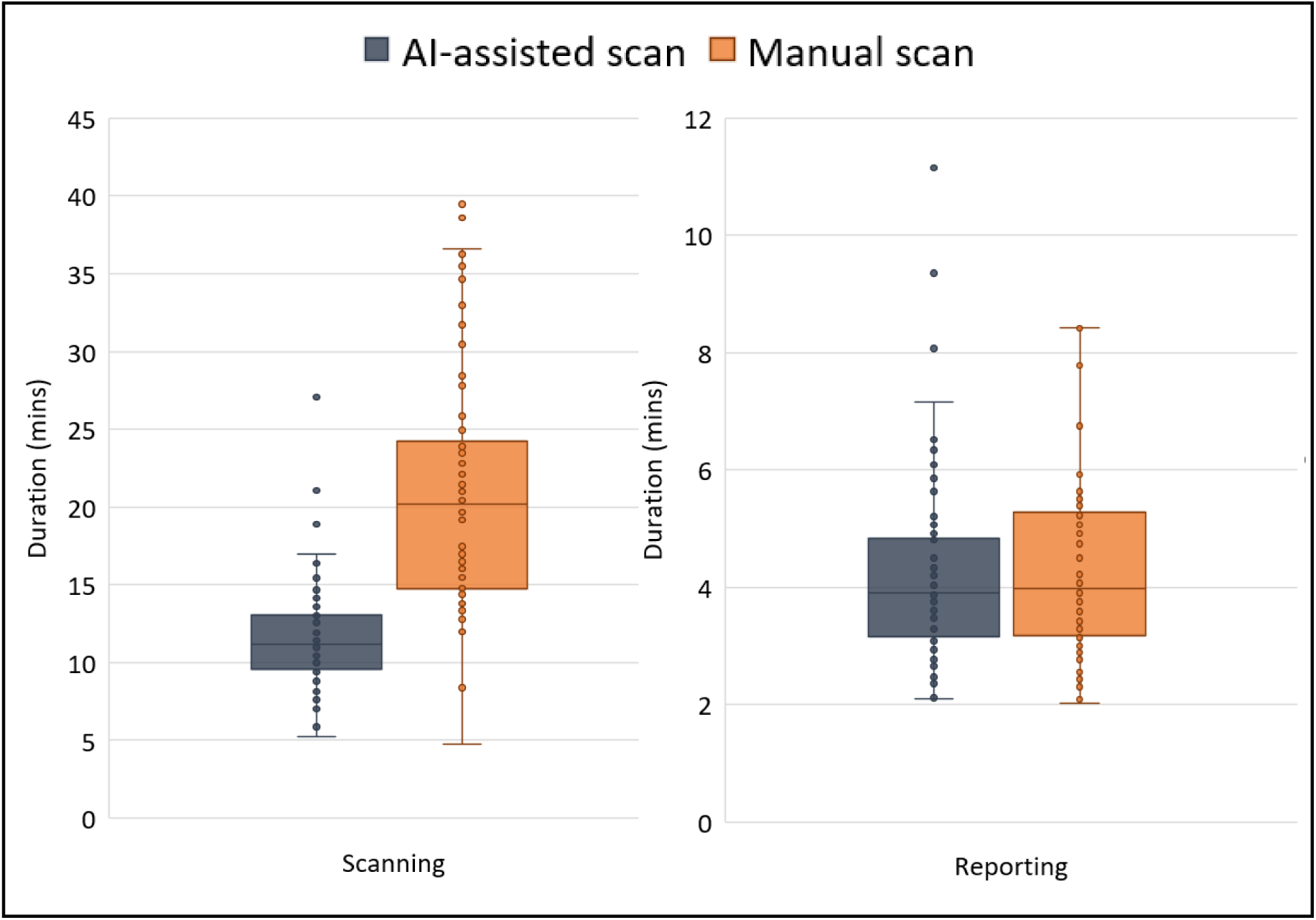
duration of scanning and reporting by both methods.

**Table 7:** scan and reporting durations. Data are medians (IQR). * Wilcoxon signed rank test.

|  | AI-assisted scan | Manual scan | P value* |
| --- | --- | --- | --- |
| Scan duration (min) | 11.4 (3.7) | 19.7 (9.6) | <0.001 |
| Report duration (min) | 3.9 (1.7) | 4.0 (2.2) | 0.335 |
| Combined scan and report duration (min) | 15.6 (3.9) | 24.1 (10.1) | <0.001 |

The cognitive load of the sonographers was compared between the two groups using both the unweighted NASA-TLX scale and the Paas scale. By both metrics, the sonographers in the AI-assisted scan group reported lower cognitive load than those in the manual scan group (NASA-TLX median score 35.3 vs 46.5 respectively, p <0.001, Paas scale 5 vs 6, p = 0.004). This is shown in Figure 6.

**Figure 6:**
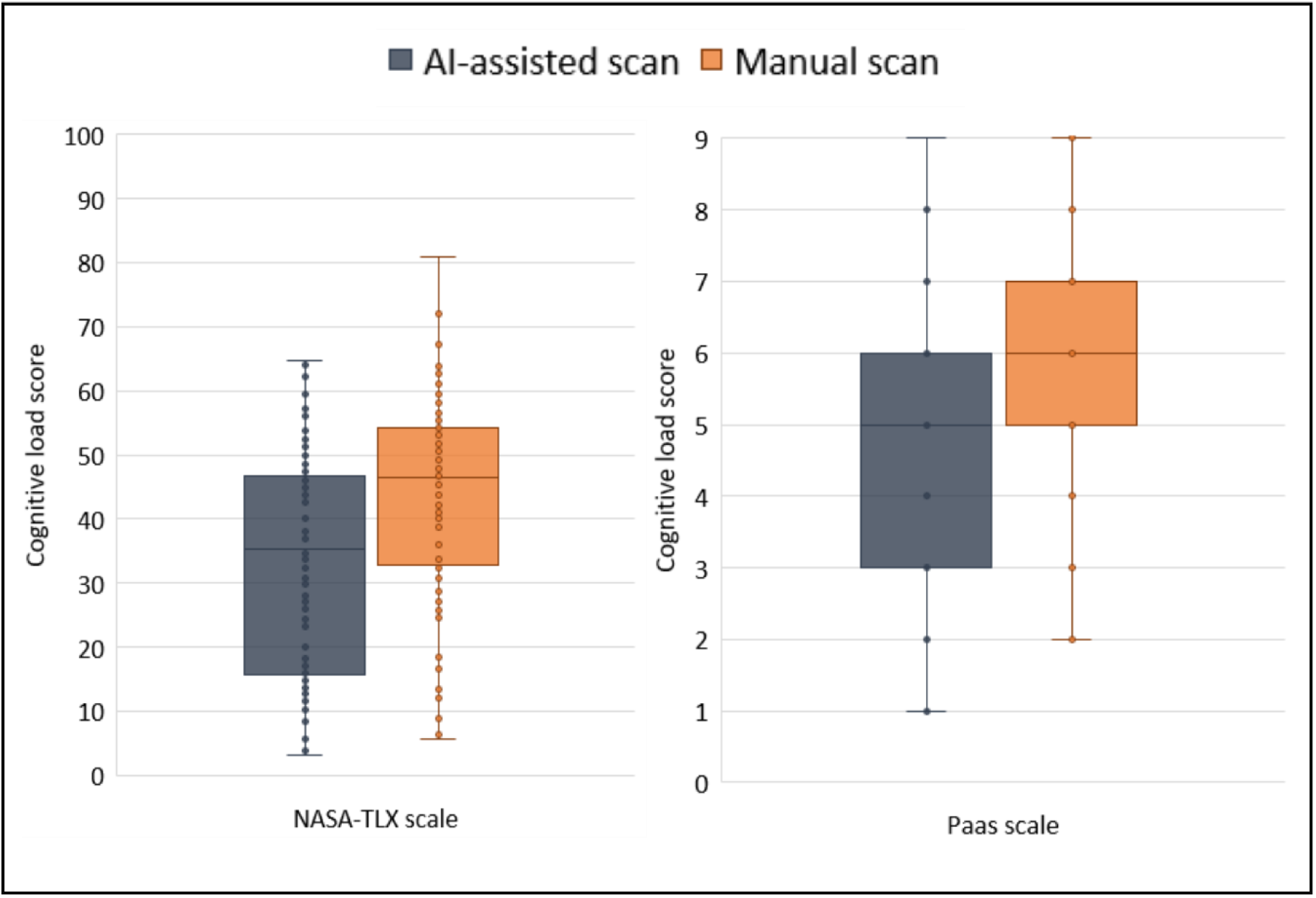
cognitive load of the sonographers compared between the two scanning methods.

Figure 7 shows the results for the biometric measurements. The repeatability of the AI measurements (column B) was superior to that of the manual method (column A). We identified a systematic bias in the reproducibility between AI and manual measurements, from -0.26 (abdominal circumference) to +4.94 mm (head circumference), shown in column C. We did not measure reproducibility between two different human observers, but this has been measured previously^23^ and is shown in red in column D (in this column the systematic bias has been removed by subtracting the mean difference, to allow direct comparison of the random error between the two groups). The random error seen when comparing AI to manual measurements was less than the random error seen between two humans.

**Figure 7:**
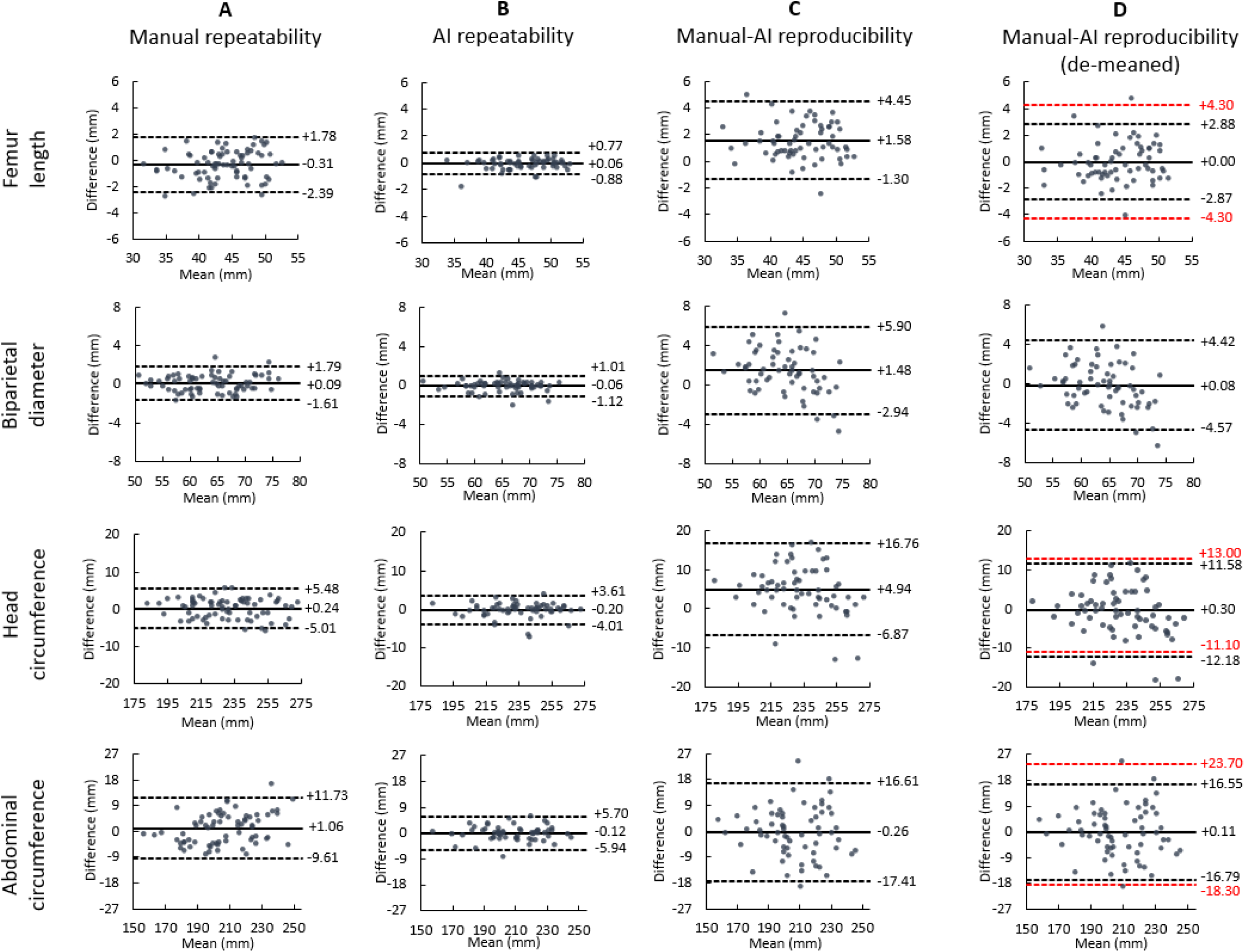
Bland-Altman plots for the four biometric measurements. Solid line: mean difference; dashed lines: the upper and lower 95% limits of agreement; A: the manual measurement repeatability, based on the three measurements taken during the manual scan; B: the repeatability of the AI measurement; C: reproducibility between the final chosen manual measurement with the measurement from the AI-assisted scan; D: as C but with the mean AI-manual difference subtracted from the AI value, to show only random and not systematic error, meaning that human interobserver variability (taken from a previous publication by Sarris et al^23^ can be directly compared (shown in red).

The quality of the images was assessed using two metrics: whether the image was “clinically acceptable” or not, and an overall quality score defined by specific parameters for each image plane (both defined in supplementary information 4). A new automatic quality assessment AI model became available during the course of the trial. This was not implemented live, but rather assessed retrospectively by running all images saved during each AI-assisted scan through the model, displaying the top 8 images in terms of assessed quality, and manually selecting the highest quality image.

The proportion of images for each method deemed clinically acceptable are shown in Table 8. Based on the AI models used live in the actual trial, image quality was significantly lower for the AI-acquired images compared to the manual scans. After the retrospective use of new AI models to automatically select the highest quality image, there was a significant improvement in image quality in many planes, meaning that for eight of the 13 planes there was no significant difference between the manually and AI-acquired images. However, for five planes (left ventricular outflow tract, brain transventricular, coronal lips, sagittal spine, and transverse kidneys), the manual scan still resulted in a higher proportion of clinically acceptable images being saved.

**Table 8:** image quality as expressed as proportion deemed clinically acceptable. ^†^quality of images selected during actual trial; **^ǂ^**quality of images selected retrospectively after use of automatic image quality assessment AI model; *p value for McNemar’s test comparing manual scans with each of the two AI methods respectively.

| Plane | Proportion of images deemed clinically acceptable (95% confidence intervals) |  |  |  |  |
| --- | --- | --- | --- | --- | --- |
|  | Manual | AI-assisted<sup>†</sup> | P value* | AI-assisted with quality model<sup>‡</sup> | P value* |
| Abdomen | 0.910 (0.815-0.966) | 0.746 (0.625-0.845) | 0.03 | 0.806 (0.691-0.892) | 0.14 |
| Four-chamber | 0.674 (0.515-0.809) | 0.465 (0.312-0.623) | 0.08 | 0.698 (0.539-0.828) | 1.00 |
| Left ventricular outflow tract | 0.674 (0.515-0.809) | 0.535 (0.377-0.688) | 0.21 | 0.442 (0.291-0.601) | 0.03 |
| Right ventricular outflow tract | 0.791 (0.640-0.900) | 0.512 (0.355-0.667) | 0.03 | 0.674 (0.515-0.809) | 0.38 |
| Three-vessel tracheal | 0.744 (0.588-0.865) | 0.512 (0.355-0.667) | 0.06 | 0.651 (0.491-0.790) | 0.45 |
| Brain transventricular | 0.925 (0.834-0.975) | 0.761 (0.641-0.857) | <0.01 | 0.791 (0.674-0.881) | 0.04 |
| Brain cerebellar | 0.806 (0.691-0.892) | 0.612 (0.485-0.729) | <0.01 | 0.806 (0.691-0.892) | 1.00 |
| Profile | 0.701 (0.557-0.807) | 0.642 (0.515-0.755) | 0.57 | 0.716 (0.593-0.820) | 1.00 |
| Coronal lips | 0.881 (0.778-0.947) | 0.333 (0.312-0.560) | 0.01 | 0.537 (0.411-0.660) | <0.01 |
| Sagittal spine | 0.791 (0.674-0.881) | 0.418 (0.298-0.545) | <0.01 | 0.388 (0.271-0.515) | <0.01 |
| Coronal spine | 0.552 (0.426-0.674) | 0.149 (0.074-0.257) | <0.01 | 0.567 (0.440-0.688) | 1 |
| Transverse kidneys | 0.806 (0.691-0.892) | 0.507 (0.382-0.632) | <0.01 | 0.567 (0.440-0.688) | <0.01 |
| Femur | 0.896 (0.797-0.957) | 0.866 (0.760-0.937) | 0.77 | 0.940 (0.854-0.983) | 0.51 |

The results of the expert quality scores for each plane are shown in Table 9. Using the AI models that automatically selected the best quality images, for two planes (left ventricular outflow tract and three vessel tracheal) the experts graded the AI images as superior to the manual images, and for three planes (brain cerebellar, coronal spine and transverse kidneys) the manual images as superior. The remaining planes were not different between methods.

**Table 9:**
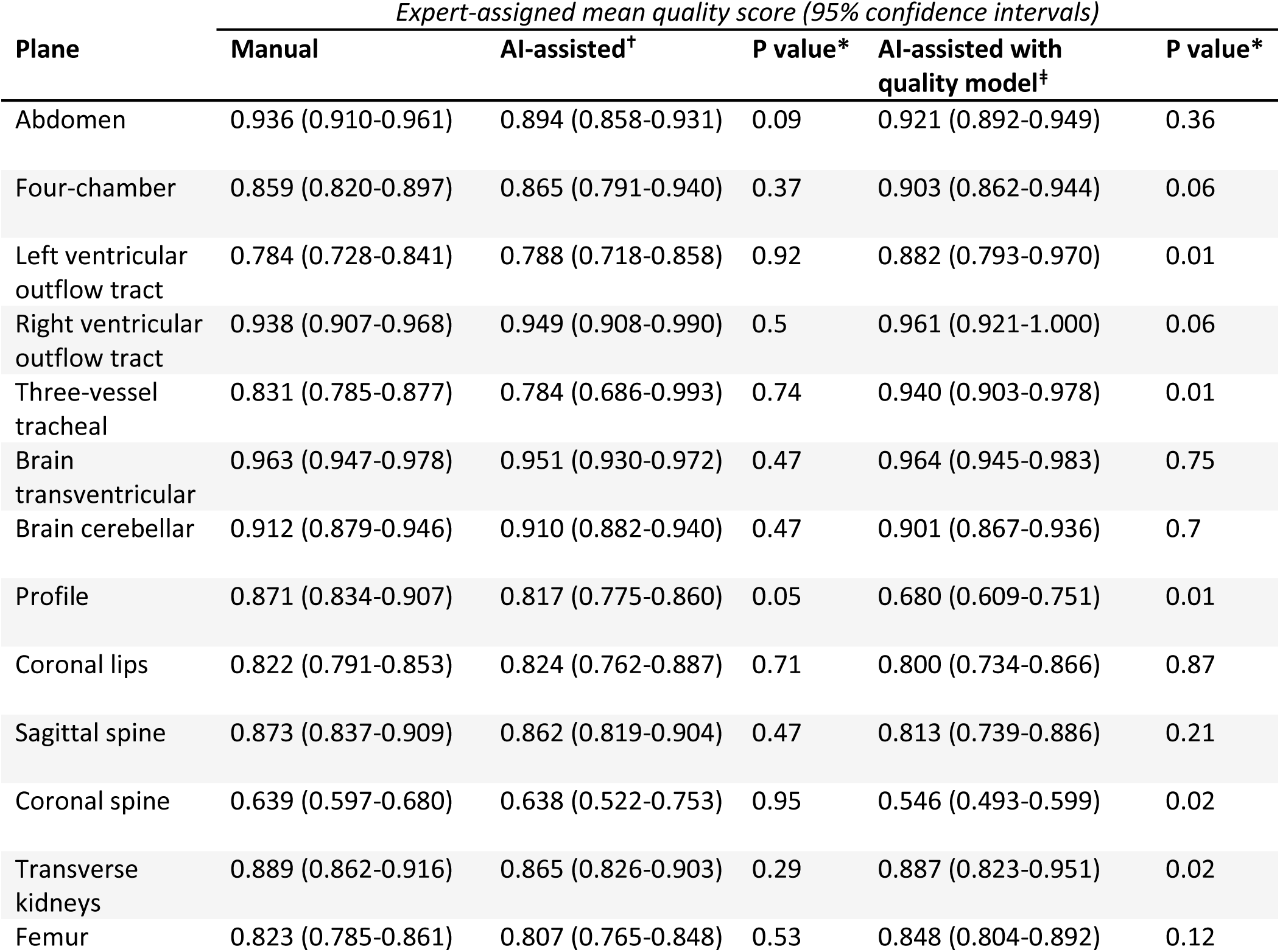
quality of images as assessed by expert scoring using specific criteria for each plane, normalised to between 0-1. Only images deemed clinically acceptable are included in analysis. ^†^quality of images selected during actual trial; **^ǂ^** quality of images selected retrospectively after use of automatic image quality assessment AI model; *p value for McNemar’s test comparing manual scans with each of the two AI methods respectively.

| Plane | Expert-assigned mean quality score (95% confidence intervals) |  |  |  |  |
| --- | --- | --- | --- | --- | --- |
|  | Manual | AI-assisted<sup>†</sup> | P value* | AI-assisted with quality model<sup>‡</sup> | P value* |
| Abdomen | 0.936 (0.910-0.961) | 0.894 (0.858-0.931) | 0.09 | 0.921 (0.892-0.949) | 0.36 |
| Four-chamber | 0.859 (0.820-0.897) | 0.865 (0.791-0.940) | 0.37 | 0.903 (0.862-0.944) | 0.06 |
| Left ventricular outflow tract | 0.784 (0.728-0.841) | 0.788 (0.718-0.858) | 0.92 | 0.882 (0.793-0.970) | 0.01 |
| Right ventricular outflow tract | 0.938 (0.907-0.968) | 0.949 (0.908-0.990) | 0.5 | 0.961 (0.921-1.000) | 0.06 |
| Three-vessel tracheal | 0.831 (0.785-0.877) | 0.784 (0.686-0.993) | 0.74 | 0.940 (0.903-0.978) | 0.01 |
| Brain transventricular | 0.963 (0.947-0.978) | 0.951 (0.930-0.972) | 0.47 | 0.964 (0.945-0.983) | 0.75 |
| Brain cerebellar | 0.912 (0.879-0.946) | 0.910 (0.882-0.940) | 0.47 | 0.901 (0.867-0.936) | 0.7 |
| Profile | 0.871 (0.834-0.907) | 0.817 (0.775-0.860) | 0.05 | 0.680 (0.609-0.751) | 0.01 |
| Coronal lips | 0.822 (0.791-0.853) | 0.824 (0.762-0.887) | 0.71 | 0.800 (0.734-0.866) | 0.87 |
| Sagittal spine | 0.873 (0.837-0.909) | 0.862 (0.819-0.904) | 0.47 | 0.813 (0.739-0.886) | 0.21 |
| Coronal spine | 0.639 (0.597-0.680) | 0.638 (0.522-0.753) | 0.95 | 0.546 (0.493-0.599) | 0.02 |
| Transverse kidneys | 0.889 (0.862-0.916) | 0.865 (0.826-0.903) | 0.29 | 0.887 (0.823-0.951) | 0.02 |
| Femur | 0.823 (0.785-0.861) | 0.807 (0.765-0.848) | 0.53 | 0.848 (0.804-0.892) | 0.12 |

## Discussion

This is the first randomised controlled trial assessing the use of AI to assist in fetal ultrasound screening, including both normal and abnormal fetuses. We have shown that use of AI can significantly reduce scan duration and sonographer cognitive load, whilst maintaining the quality of the scan in terms of disease detection. Automatically measured fetal biometric measurements were more repeatable and reproducible compared to human manual measurements. Image quality for some planes was initially inferior using the AI tools, but this was partially ameliorated by the retrospective use of AI models to automatically select the highest quality images (although further work is needed to improve this for some planes). We used a trial design that allowed for a reasonable sample size through enrichment with fetuses affected by CHD, and controlled for variation in both sonographers (through the use of randomisation) and pregnant participants (as all were scanned using both methods).

Previous work in the field has focused on assessment of algorithm performance using retrospective curated test datasets, which may not fully reflect the performance achieved in a real clinical environment. ^9–14^ A small pilot study by our group has previously suggested a significant time saving with the use of AI, and the present study expands on this by including fetuses with known structural malformations (so that diagnostic performance of the human-AI team could be assessed), and involving a large cohort of randomised and blinded sonographers.^9^ These results are encouraging, and suggest that a real clinical benefit may be offered if AI is integrated into current fetal ultrasound screening programmes.

We have shown that on average sonographers with AI assistance saved around 42% of the scan duration, which is time that could be then directed towards other tasks to improve the overall scan experience, such as communicating with the patient or spending more time imaging a particular anatomical area of concern. There may also be important health economic benefits to shorter scan durations, due to a reduction in scan cost.

Our hypothesis was that (secondary to a reduction in cognitive load) the diagnostic performance of the scan would be improved by AI assistance. The improvement in screening sensitivity for CHD did not reach statistical significance, but there was a small improvement seen in specificity, if considering CHD only. Considering all fetal malformations, there was no difference in specificity between the two methods. As we have previously demonstrated, specificity is extremely important when considering the introduction of AI to screening programmes, to avoid overwhelming downstream specialist services with false positive referrals.^24^ The fact that specificity remains robustly high in the AI-assisted group offers reassurance that AI tools may prevent this issue, potentially reducing false positive referrals for CHD.

The analysis of biometric measurements is also encouraging. We have shown that the automatically measured biometrics were both more repeatable, and with less random error, compared to another manual measurements by a different sonographer. We did identify a difference between manual and AI measurements, but without a gold standard it is not clear which is the more accurate. It would be relatively trivial to convert the AI measurement to an equivalent of the manual measurement by subtracting the detected difference, if that were desirable. The AI measurements were based on tens or hundreds of measurements per scan using a Bayesian approach^18^, rather than the traditional approach of measuring just three times (or in many cases, just once). This resulted in a final estimate that had a far higher repeatability compared to manual measurements, as well as having the advantage of real time feedback to the sonographer of the error range around the current estimated measurement. This method could be applied to many ultrasound-based measurements, even beyond obstetrics. By reducing random human error, we can obtain measurements that are precise, even if the scan is conducted by a different operator. Such measurements are often extremely clinically important, and by reducing variability we can be more confident about thresholds used to instigate or monitor treatments.

Even though AI-assisted sonographers were using a novel system after only a very short training period, they still recorded significantly lower cognitive load scores by two different metrics compared to the manual group. Cognitive load is a concept describing how mentally challenging a given task is, and reducing it - by taking over specific mundane and/or distracting tasks - is a potential mechanism by which the human-AI team performance might exceed that of humans alone.^25^ The combination of reduced cognitive load and reduced scan duration shows exciting potential, and one we hope will be translated into improved fetal ultrasound screening outcomes.

The results for image quality are mixed. Our initial results for image quality showed that images were graded as lower quality when AI-acquired compared to manually acquired images. However, when we utilised an improved AI model retrospectively to automatically select the highest quality images from the same recorded examinations, this problem was solved for many of the planes. This indicates that the problem for these planes was not that high quality images were not saved, it was that the candidate images presented to the sonographers were initially not the subjectively “best” ones out of all the saved images. Our current image quality models have performed well, but some image planes still require further improvement - probably via an enlargement of the training set with further labelled images - to match the quality of manually saved images. These quality models could be easily integrated into the overall clinical tool and used in real time for future studies.

The main limitation of this trial was that we conducted research ultrasound scans, performed in addition to the standard clinical pathway. The sonographers were self-selected, and as such may not reflect the overall population of sonographers, either in terms of professional background, or skill level. Although they were blinded to the CHD status of each fetus, and even that CHD was the focus of the study, they were aware that a potential malformation was present. Given each sonographer only scanned a maximum of three participants, they may have been more cautious or thorough compared to their usual clinical practice. Because of the current limitations of fetal ultrasound screening, we could not recruit pregnant participants from the standard screening population as we would not have access to a reliable ground truth. For this reason, we recruited participants who had undergone detailed fetal echocardiography, a procedure that in expert hands has a much higher sensitivity and specificity than standard ultrasound screening.^26^ This means that by definition we only included cases of CHD that had already been detected. How well AI tool assistance works in an unselected population has not yet been assessed. We also used a single model of ultrasound machine, meaning that potential domain-shift problems that may be encountered in clinical use have not yet been fully explored.

We have not addressed some broader concerns regarding medical AI in this trial, such as the potential for workforce deskilling by the automation of specific tasks. This is an important issue and will need to be addressed if AI is to be translated to the clinical environment, perhaps by ensuring sonographers still undertake some manual scans intermittently. However, some other broader concerns such as inattention to anomalies secondary to “automation bias” have been addressed in this study, and our findings are reassuring on this front. Many risks of AI are at least partially mitigated by ensuring that the AI tool and human operator work in partnership, and that the human (in this case the sonographer) always retains complete control over the final interpretation of the scan findings.

Translation of this study to real-world screening-level population will be key to fully explore the utility of AI tools. Given the prevalence of congenital anomalies in the general population this will likely require a large multi-site trial, recruiting participants as they undergo their routine clinical screening ultrasound. Previous work has also explored the use of AI to directly detect anomalies such as CHD on ultrasound images.^24,27–29^ The addition of such models to our current AI tool may further improve overall scan performance, but this does not come without risk, and needs to be carefully assessed. However, such model ensembles may be a powerful way of improving detection of fetuses with congenital malformations, and will be the focus of our future work.

In summary, we have demonstrated that AI-assistance for fetal ultrasound screening is safe and effectively reduces scan duration and cognitive load, without a reduction in diagnostic performance. This is one of the relatively few randomised prospective controlled trials of AI in medicine and raises the exciting prospect of future human-AI collaboration in this field.

## Acknowledgements

Firstly, we would like to thank all the pregnant and sonographer participants of this trial, who gave up their time to try and improve fetal ultrasound screening. We would also like to thank the nursing and medical staff of the Evelina Children’s Hospital Fetal Cardiology Unit, for their help in recruiting the participants for this study. Finally, we would like to thank Caitlin Giles and Abigail Adeosun, along with the staff of the St Thomas’ Hospital Clinical Research Facility for the smooth running of the trial.

Figure 1 and 3 were created using images from Flaticon.com.

The study was funded by an NIHR doctoral fellowship (NIHR301448) and was supported by grants from the Wellcome Trust (IEH Award, 102431), by core funding from the Wellcome Trust/EPSRC Centre for Medical Engineering (WT203148/Z/16/Z), and the London AI Centre for Value Based Healthcare via funding from the Office for Life Sciences.

BK received funding by the ERC - project MIA-NORMAL 101083647

## Data availability

Patient-level imaging data from the trial, and the imaging data used to train the AI models are not available for sharing due to ethical restrictions. The study protocol, patient information sheet, and example consent forms are available on request. The code used to train the AI models is available on request, but the model weights used in the trial are not available.

## Supplementary information 1: development of AI models

### Dataset

The data used to train the AI models used for this paper were collected as part of the iFIND project^30^. The entire videos of 9,739 routine fetal anomaly ultrasound examinations were recorded prospectively as they were performed. This dataset was named iFIND1. These examinations were conducted by over 100 professional sonographers on identical ultrasound machines (GE Voluson E8) between 2015 and 2020 in a single central London teaching hospital. During these examinations, sonographers acquired and labelled standard plane images and measured biometrics in real time. To best reflect the screening population, the scans were not selected for normality. Due to operator error, technical glitches, and patient withdrawal of consent, not all these scans could be used in the dataset, meaning 7309 video recordings of prenatal ultrasound scans were used for this study. The demographic details for the dataset are shown in Table 10

**Table 10:**
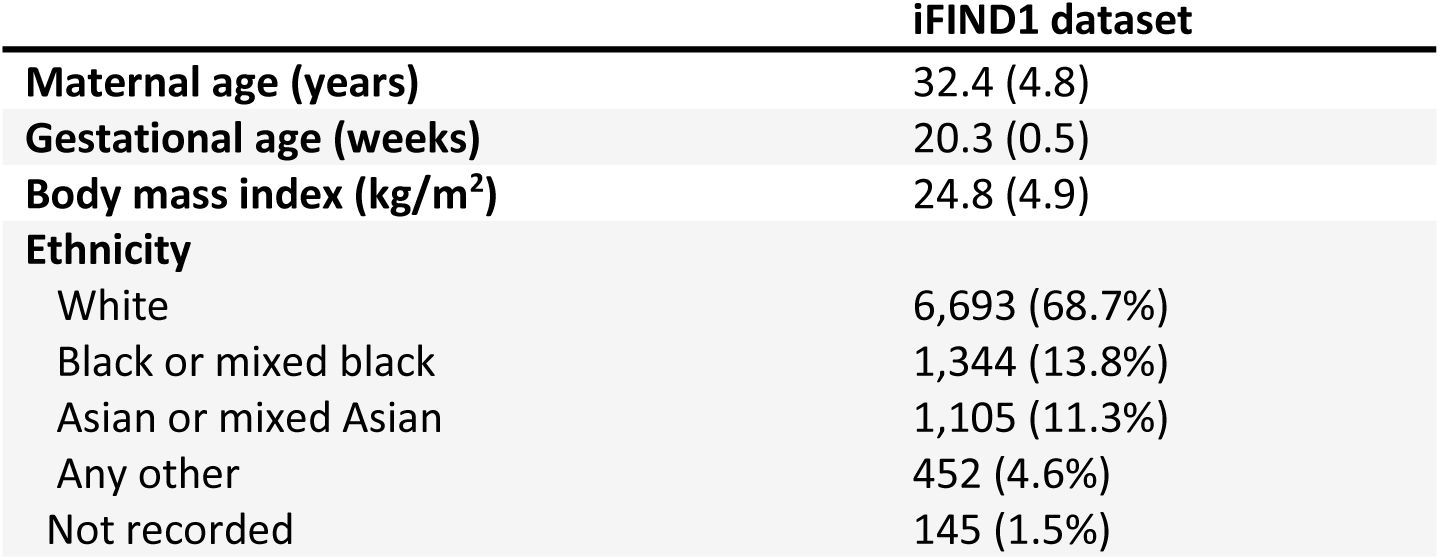
demographic information on iFIND1 dataset.

|  | **iFIND1 dataset** |
| --- | --- |
| **Maternal age (years)** | 32.4 (4.8) |
| **Gestational age (weeks)** | 20.3 (0.5) |
| **Body mass index (kg/m<sup>2</sup>)** | 24.8 (4.9) |
| **Ethnicity** |  |
| White | 6,693 (68.7%) |
| Black or mixed black | 1,344 (13.8%) |
| Asian or mixed Asian | 1,105 (11.3%) |
| Any other | 452 (4.6%) |
| Not recorded | 145 (1.5%) |

To maintain patient anonymity, personal information was removed from the scan recordings and each scan was labelled with a numerical study ID, numbered sequentially. We used study IDs to create consistent training/validation/test splits across the dataset: the trailing digit determined which split each scan assigned. Scans with a study ID with a trailing digit between 0-5 were used to train our models, 6-7 were used for validation during training, and 8-9 were held aside for testing and not used at all during training of any of our models, giving a test/validation/test split of 60%/20%/20% respectively.

To find plane labels, we automatically detected any pauses and freezes in the videos, then used OCR software to extract any text labels that were added to the frames. The text labels were then associated with standard planes, and manually checked for consistency.

Biometric annotations were also performed by sonographers in real time by placing calliper markers on the structure of interest. We extracted the locations of those callipers within the image to train our biometric networks.

### Standard plane classification

We used the data and annotations described above to train an AI model to classify for fetal standard planes. We used the Sononet architecture as previously described^31^ and trained it on the training labels described above, as well as “background” frames corresponding to no standard plane. This classifier obtained >90% top-1 classification accuracy in the iFIND test dataset (taken from scans with trailing digit 8-9). We named this model SonoNEXT.

### Image clustering and final image selection

Because of the nature of video (20-30 images per second), there will be many valid standard view images during a scan. During live scanning SonoNEXT operates on an image-per-image basis, and thus saves multiple, often several hundreds valid images for each plane. However, only one image is required for archiving purposes. This single image can be selected by the human operator, but it is not feasible to look through hundreds of images to do this, hence some sorting or clustering step is required to reduce the number of candidate images for the human to choose from.

To do this, at the end of the scan, for each plane all valid images were sorted into 20 clusters, using a K-means clustering algorithm. This groups images based on chronological timestamp for when the image was acquired, the output feature vector of SonoNEXT, recorded pixel size, and an image sharpness metric. The image that represents the centroid of each cluster was then selected, giving 20 images. These 20 images were then ranked, based on timestamp (later images being ranked higher), pixel size (more zoomed images ranked higher), image sharpness, and a metric defining how centred the region of interest was (defined by the SonoNEXT feature attention heatmap relative to the entire image). This ranking of images was used during the “image review stage”, after the scan is complete.

### Image quality

After the trial had completed, it became apparent that the images presented to the sonographer after the clustering step described above were anatomically correct but of a lower quality than the manually-acquired images. We felt that this was because the image clustering step did not fully take into account expert-perceived image quality, and so there were likely many images of better quality that were not being presented to the sonographers, meaning an inferior quality image was ultimately chosen as the ‘best’ image. Therefore, we devised a method to automatically regress a quality score for each image of each standard plane and presented the image with the highest predicted quality. We trained a CNN to predict the clinical quality of images classified as valid standard plane. This method could then be incorporated into the clustering step for future versions of the AI tool.

### Quality dataset labelling

We generated training datasets for each of our models from the outputs of the SonoNEXT standard- plane detection model. We randomly selected a single image for each standard plane from the test set of the SonoNEXT model (n=1457 subjects). There was no pre-selection step: the random selection intentionally included SonoNEXT misclassifications and poor-quality views to be representative of the images that would be passed to the quality models during scanning.

A set of objective clinical criteria were devised to evaluate the quality of each standard plane, decided by a committee of clinical experts. In addition to these criteria, we also asked labellers whether each image was overall “clinically acceptable”. We also asked them to rate the subjective quality of each image on a scale of 1-10.

### Quality model training

We trained models using the same SonoNEXT architecture that we used for plane-classification to evaluate the quality of images. We modified the output layer to predict each feature that had been labelled for this standard plane, turning the network into a multitask classification network. We used cross-entropy as the loss function in training.

### Image scoring and presentation

After each image for a standard plane was processed through our quality models, we selected 9 images to present to the sonographer to make a manual selection of the best one. To select the images to present, we calculated two scores for each image, which we called the quality score and the diversity score.

The quality score is a single score using a composite measure derived from the output vector for each image. Not all of the features labelled and predicted by the network are equally important: some should be given a higher weight when determining an image’s quality. To quantify the importance of each feature, we calculated a multivariate logistic regression to predict whether each image was rated “clinically acceptable” based on all the other labelled features. This had a predictive accuracy of >95% for all standard planes. The quality score was therefore a weighted sum of the predicted features, weighted by the regressed weights associated with that plane. Each image had a quality score calculated in this way.

In a single scan, the images with the highest quality score are usually visually very similar to each other and are often consecutive frames acquired very close to each other. Presenting the images with the highest quality score for the sonographer to select from would only offer a very restricted choice. To address this, we added a diversity score, designed to be higher for images that are more different from those already presented.

We defined the diversity score *D_i_* for each sample x_i_ as the lowest Euclidean norm between its output vector from the quality CNN and that of images that have already been selected

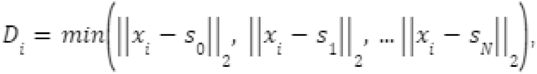

where s_0_, s_1_, … s_N_ are the CNN output vectors of the N previously selected images. An image identical to one that has already been presented will therefore have a diversity score of 0, while all other images will have a score inversely proportional to their similarity to the most similar previously selected image.

This is an iterative process: the diversity score is initially undefined. The first image to be chosen is therefore simply the one with the highest quality score. After this, the diversity score can be calculated for all images with reference to the first selected image. This score can be updated after each successive image is selected to be presented to the sonographer.

The images presented to the sonographer were chosen based on a weighted sum of the quality score and the diversity score. The choice of weight is important: a very low weight for the diversity score is likely to result in many very similar images being presented for selection, raising the chances that none of them are clinically acceptable. A very high weight for the diversity score will prioritise images that are very different from each other, but this will often include misclassifications and poor-quality views. We empirically decided to use a weight of 0.05 for the diversity score after manual grid search of an optimal value for this hyper-parameter.

### Biometrics

Additionally, we trained AI models to measure the four key fetal growth biometrics of head circumference (HC), biparietal diameter (BPD), abdominal circumference (AC) and femur length (FL). We used the coordinates of the calliper locations to generate heatmaps consisting of Gaussian kernels centred at the calliper locations. We then trained a segmentation network using the U-Net architecture^32^ to predict the heatmap for every image in the training set. We trained a separate segmentation network for each of HC, AC, and FL. BPD was not measured independently: we measured the minor axis of the derived HC ellipse to calculate it. We named these models BiometricNet-Head, BiometricNet-Abdo, and BiometricNet-Femur.

There are many frames in each scan in which biometrics can be measured. With the methods described above it is possible to extract these measurements in all frames in which the biometric is visible, resulting in hundreds or thousands of measurements per biometric. These are likely to be slightly different from each other, as the views are themselves somewhat different.

We used a Bayesian framework to aggregate all measurements into a global estimate of each biometric, which was updated in real time as more frames were visualised. With every new frame, we updated an internal model of the expected distribution of measurements. This allowed us to extract a central estimate of each biometric as well as a credible interval in which the biometric can be expected to lie. The live estimates and credible intervals were displayed to the operator during scanning. This approach is explained in more detail in Venturini *et al*.^18^

## Supplementary information 2: development of clinical tool

The clinical tool used in this study was developed in three parts:

1. Backend software application
2. Frontend tablet application
3. Frontend reporting application

### Backend software application

This was developed in Python and served four purposes:

1. Read and process a live stream of ultrasound images. Images are read from a video capture card, pre-processed and passed to our SonoNEXT AI model. Outputs of this model are then used to determine which (if any) biometry machine learning model to pass to. The image is also passed to a pixel size estimator class. The outputs of the biometry and pixel size estimator are combined to give measurements in millimeters. We use this measurement to update the current estimate of each biometric.
2. Convert the analysis to JSON and stream over websocket to the frontend application during live scanning. The outputs of Step 1 are converted to JSON format and streamed over WebSocket technology to the frontend application for display.
3. Save the images and analysis to a local database (MongoDB). Every image is converted to base64 jpeg encoding, paired with the JSON from step 2 and saved to a local NoSQL database solution.
4. Expose a REST API to query the local database during reporting on both the tablet and reporting applications. The main functions are to:

- READ images from a scan for each SonoNEXT classification, and cluster and rank these to give 9 candidate images per classification.
- WRITE a report object for the patient to the database.
- READ the report object for a patient.
- UPDATE the report object for a patient.

### Frontend tablet application

This was developed in Javascript / HTML / CSS and served the following purposes through the following UI screens:

1. ‘New Scan’ Screen: Input patient information to start a new scan including ID, sonographer ID, EDD (+GA is calculated automatically), the ultrasound machine being used and whether the scan was to be an ‘AI assisted’ scan or not. (If not ‘AI assisted’ then the frontend application would be blank until the end of the scan in order to capture data in the background but not display this to the sonographer).
2. ‘Live Scan’ Screen: The application receives live stream of image analysis from the backend via websocket and displays the following:

- A ‘Plane’ card for each standard plane: Name and progress bar displaying how many images have been collected and classified as that plane.
- A ‘Biometry’ card for each biometric: Name, current estimate in millimeters, place on centile for that GA and indicator of statistical validity for that measurement’s current estimate.
3. ‘Report’ Screen: Once scanning is complete, the sonographer clicks a button to navigate to the reporting stage of the scan (in non-AI assisted scanning this button ends the scan and returns to screen 1.). Here the sonographer is shown the patient details, a list (with images) of the standard plane ‘candidates’ and the biometrics (with images with overlays) collected during the scan. The sonographer can click of a standard plane to choose between up to 9 alternative candidate images to save in the report instead of the initially chosen image. Finally, the sonographer indicates if the scan was ‘Normal’ or ‘Needs further information’ before finishing the scan and returning to screen 1.

### Frontend reporting application

The fronted reporting application was developed in Javascript / HTML / CSS and served the following purposed through the following UI screens:

1. ‘Find report’ screen: The UI displays a list of recently completed scans and the sonographer selects the scan they wish to complete the report for.
2. ‘Report’ screen: The UI displays the fields and biometrics collected during the scan or leaves them blank for completion if it was not an ‘AI assisted’ scan. The sonographer completes the remaining items on the report on this screen.
3. ‘Preview’ screen: The UI displays a preview of the completed report. It is then uploaded to the local database. Return to screen 1.

### Technology stack

Frontend Tablet: Javascript, HTML, CSS, VueJS, CapacitorJS, NodeJS

Frontend Report: Javascript, HTML, CSS, VueJS, NodeJS

Backend Software: Python, FastAPI, WebSocket, Sklearn, ONNX, OpenCV, Numpy

### Models used

The final models used in this study were:

- SonoNext v1.0

- BiometricNet-Head v1.0

- BiometricNet-Abdo v1.0

- BiometricNet-Femur v1.0

## Supplementary information 3: scan protocols

### Protocol for AI-assisted anomaly ultrasound scan

- Please perform a modified ultrasound anomaly scan, using the table below as a guide, taking a maximum of 30 minutes.
- Please observe the 6 main anatomical areas:

`o` an example image for the 13 planes identified below will be saved automatically if you scan it.
`o` the 4 biometric measurements below will be measured automatically and displayed to you.
- Please do not save additional images or measure additional biometric measurements.
- Do not use colour Doppler or spectral Doppler.
- Do not assess the cervix, amniotic fluid volume, placenta, cord vessels, or presentation.

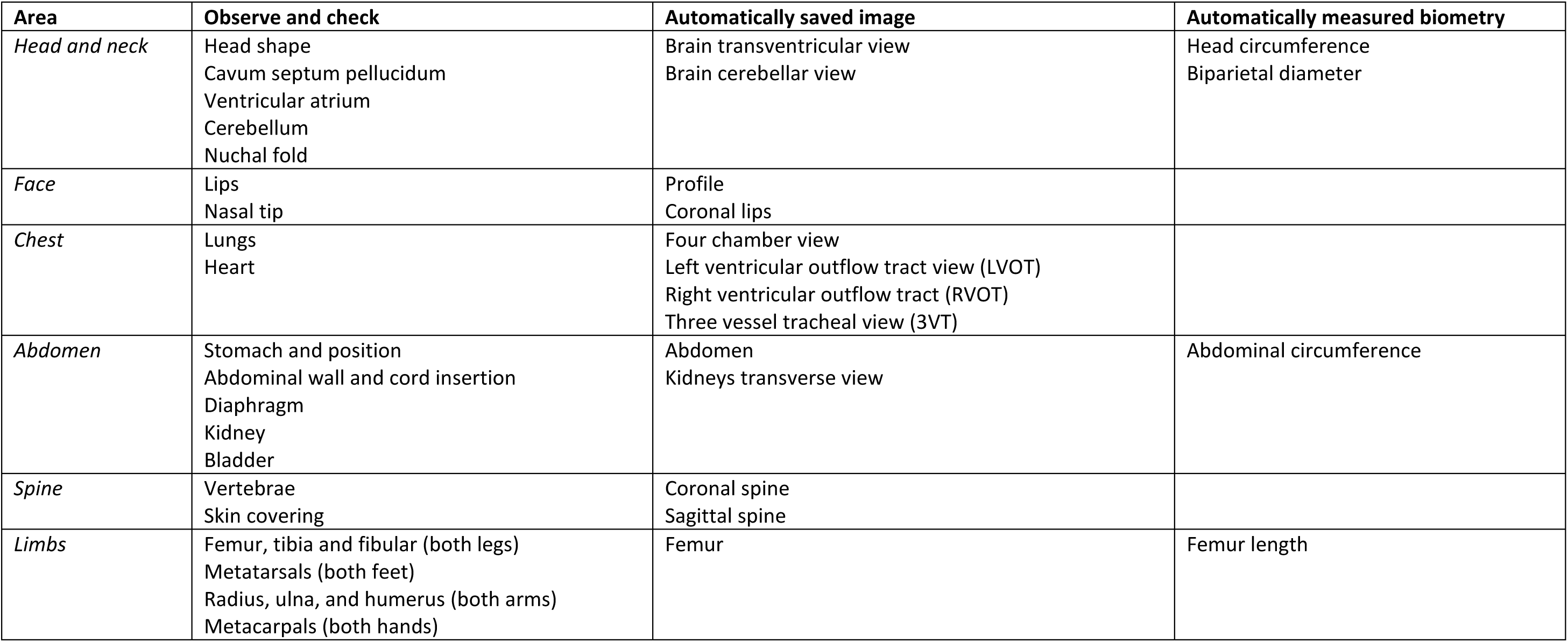

### Protocol for manual anomaly ultrasound scan

- Perform a modified ultrasound anomaly scan, using the table below as a guide, taking a maximum of 30 minutes.
- Observe the 6 main anatomical areas:

`o` save an example image for the 13 planes identified below.
`o` measure the 4 biometric measurements (measure each on three separate images and choose best).
- Do not save additional images or measure additional biometric measurements.
- Do not use colour Doppler or spectral Doppler.

| Area | Observe and check | Save image | Measure biometry |
| --- | --- | --- | --- |
| *Head and neck* | Head shape<br>Cavum septum pellucidum<br>Ventricular atrium<br>Cerebellum<br>Nuchal fold | 1. Brain transventricular view<br>2. Brain cerebellar view | 1. Head circumference<br>2. Biparietal diameter |
| *Face* | Lips<br>Nasal tip | 3. Profile<br>4. Coronal lips |  |
| *Chest* | Lungs<br>Heart | 5. Four chamber view<br>6. Left ventricular outflow tract view (LVOT)<br>7. Right ventricular outflow tract (RVOT)<br>8. Three vessel tracheal view (3VT) |  |
| *Abdomen* | Stomach and position<br>Abdominal wall and cord insertion<br>Diaphragm<br>Kidney<br>Bladder | 9. Abdomen<br>10. Kidneys transverse view | 3. Abdominal circumference |
| *Spine* | Vertebrae<br>Skin covering | 11. Coronal spine<br>12. Sagittal spine |  |
| *Limbs* | Femur, tibia and fibular (both legs)<br>Metatarsals (both feet)<br>Radius, ulna, and humerus (both arms)<br>Metacarpals (both hands) | 13. Femur | 4. Femur length |

Do not assess the cervix, amniotic fluid volume, placenta, cord vessels, or presentation.

## Supplementary information 4: image quality scoring

To assess the saved images in terms of quality, a committee comprising of six experts (consultants in fetal medicine with at least 10 years’ experience in obstetrics). These experts had Two metrics were decided upon, a binary score to define if the image was “clinically acceptable” or not, and a continuous quality score based on specified criteria for each plane.

For the clinical acceptability score, we defined “clinically acceptable” as being of *sufficient quality to reasonably assess relevant sonographic signs that correspond to the 11 FASP conditions that are screened for at the anomaly ultrasound scan*. The 11 conditions are defined in the FASP guidelines^7^. To score each image, we asked the experts to consider whether the image was of sufficient quality to allow reasonable assessment of the signs and related conditions shown in Table 11 below. The condition did not necessarily need to be ruled out, as that is not possible for some conditions with a single image plane, but should have allowed reasonable assessment, and give the impression that the sonographer would have been able to exclude the condition during live scanning.

**Table 11:** FASP conditions relevant to each image plane, and the relevant signs used to consider such diagnoses.

| Plane | FASP conditions to assess for (relevant sign(s) in brackets) |
| --- | --- |
| **4CH** | - **Major CHD** seen in 4CH view such as hypoplastic left heart or AVSD (abnormal cardiac axis, unbalanced ventricles, septal defects, abnormal offset of AV valves)<br>- **Congenital diaphragmatic hernia** (bowel / liver / stomach in thorax) |
| **LVOT** | - **Major CHD** seen in LVOT view such as tetralogy of Fallot (lack of continuity of outlet septum with aortic valve) |
| **RVOT** | - **Major CHD** seen in RVOT view such as tetralogy of Fallot (unbalanced great arteries) |
| **3VT** | - **Major CHD** seen in 3VT view such as TGA (unbalanced great vessels, abnormal orientation of vessels) |
| **Abdomen** | - **Gastroschisis** (defect in abdominal wall)<br>- **Exomphalos** (defect in abdominal wall)<br>- **Congenital diaphragmatic hernia** (absent stomach)<br>- **Bilateral renal agenesis** (absent stomach)<br>*Should also allow accurate measurement of abdominal circumference* |
| **Kidneys** | - **Bilateral renal agenesis** (kidneys not seen) |
| **Brain TV** | - **Anencephaly** (absence of calvarial bones)<br>- **Spina bifida** (lemon shaped head)<br>- **T13/18** (absent CSP / ventriculomegaly / choroid plexus cysts / strawberry shaped skull / holoprosencephaly / microcephaly)<br>- **Lethal skeletal dysplasia** (clover leaf skull)<br>*Should also allow accurate measurement of head circumference and BPD* |
| **Brain CB** | - **Spina bifida** (banana shaped cerebellum)<br>- **T13/18** (posterior fossa anomalies / nuchal thickening or cystic hygroma / cerebellar hypoplasia) |
| **Coronal spine** | - **Spina bifida** (defect in lumbosacral region, loss of normal vertebral alignment) |
| **Sagittal spine** | - **Spina bifida** (defect in lumbosacral region, possibly involving skin covering) |
| **Profile** | - **Signs of T13/18** (micrognathia)<br>- **Spina bifida** (frontal bossing) |
| **Lip** | - **Cleft lip** (defect in upper lip) |
| **Femur** | - **Lethal skeletal dysplasia** (abnormal shape / reduced echogenicity / absence of extremities)<br>*Should also allow accurate measurement of femur length* |

The second metric was an overall quality score, using between three and seven criteria per image plane which were decided upon after discussion, which were felt to be the most important factors in differentiating a good-quality image from a poor-quality image. These criteria are shown in Table 12. One point was given for each criterion, and for each image the score was summed and divided by the total number of criteria for that plane, giving a normalised quality score of between 0 – 1.

**Table 12:**
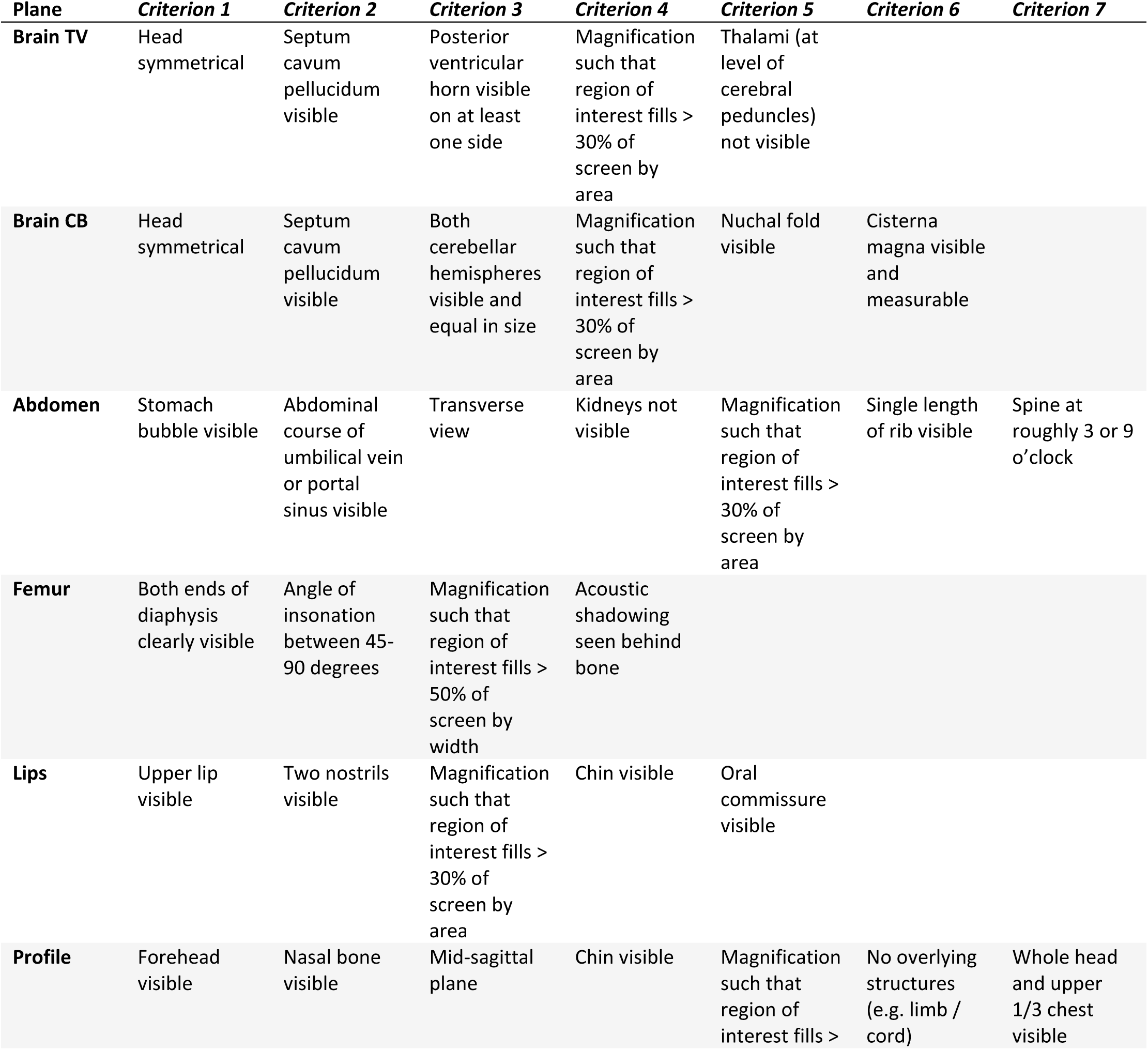
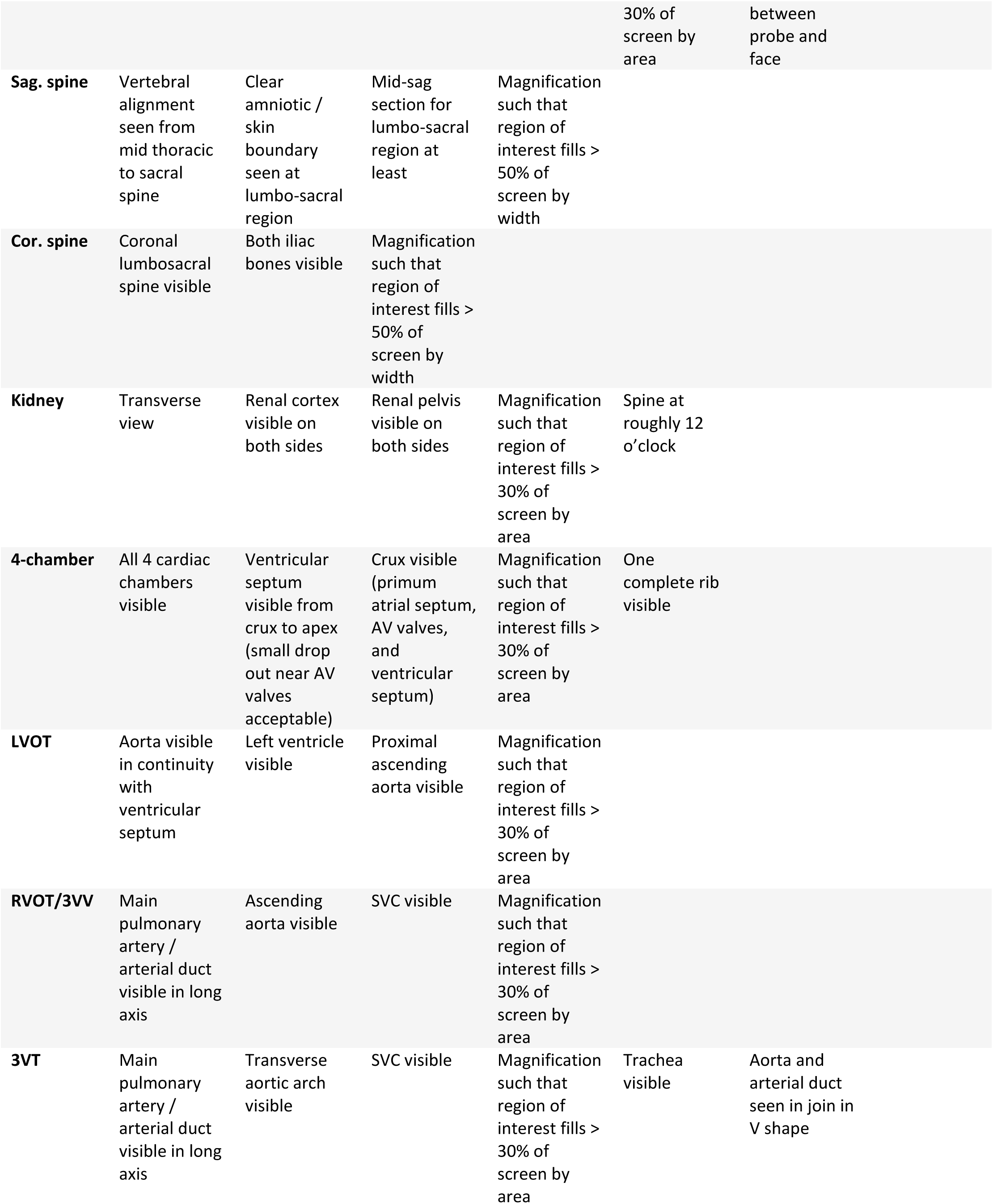
scoring criteria for each plane to give an overall quality score.

| Plane | Criterion 1 | Criterion 2 | Criterion 3 | Criterion 4 | Criterion 5 | Criterion 6 | Criterion 7 |
| --- | --- | --- | --- | --- | --- | --- | --- |
| **Brain TV** | Head symmetrical | Septum cavum pellucidum visible | Posterior ventricular horn visible on at least one side | Magnification such that region of interest fills > 30% of screen by area | Thalami (at level of cerebral peduncles) not visible |  |  |
| **Brain CB** | Head symmetrical | Septum cavum pellucidum visible | Both cerebellar hemispheres visible and equal in size | Magnification such that region of interest fills > 30% of screen by area | Nuchal fold visible | Cisterna magna visible and measurable |  |
| **Abdomen** | Stomach bubble visible | Abdominal course of umbilical vein or portal sinus visible | Transverse view | Kidneys not visible | Magnification such that region of interest fills > 30% of screen by area | Single length of rib visible | Spine at roughly 3 or 9 o'clock |
| **Femur** | Both ends of diaphysis clearly visible | Angle of insonation between 45-90 degrees | Magnification such that region of interest fills > 50% of screen by width | Acoustic shadowing seen behind bone |  |  |  |
| **Lips** | Upper lip visible | Two nostrils visible | Magnification such that region of interest fills > 30% of screen by area | Chin visible | Oral commissure visible |  |  |
| **Profile** | Forehead visible | Nasal bone visible | Mid-sagittal plane | Chin visible | Magnification such that region of interest fills > | No overlying structures (e.g. limb / cord) | Whole head and upper 1/3 chest visible |

|  |  |  |  |  |  |  |
| --- | --- | --- | --- | --- | --- | --- |
|  |  |  |  |  | 30% of screen by area | between probe and face |
| **Sag. spine** | Vertebral alignment seen from mid thoracic to sacral spine | Clear amniotic / skin boundary seen at lumbo-sacral region | Mid-sag section for lumbo-sacral region at least | Magnification such that region of interest fills > 50% of screen by width |  |  |
| **Cor. spine** | Coronal lumbosacral spine visible | Both iliac bones visible | Magnification such that region of interest fills > 50% of screen by width |  |  |  |
| **Kidney** | Transverse view | Renal cortex visible on both sides | Renal pelvis visible on both sides | Magnification such that region of interest fills > 30% of screen by area | Spine at roughly 12 o'clock |  |
| **4-chamber** | All 4 cardiac chambers visible | Ventricular septum visible from crux to apex (small drop out near AV valves acceptable) | Crux visible (primum atrial septum, AV valves, and ventricular septum) | Magnification such that region of interest fills > 30% of screen by area | One complete rib visible |  |
| **LVOT** | Aorta visible in continuity with ventricular septum | Left ventricle visible | Proximal ascending aorta visible | Magnification such that region of interest fills > 30% of screen by area |  |  |
| **RVOT/3VV** | Main pulmonary artery / arterial duct visible in long axis | Ascending aorta visible | SVC visible | Magnification such that region of interest fills > 30% of screen by area |  |  |
| **3VT** | Main pulmonary artery / arterial duct visible in long axis | Transverse aortic arch visible | SVC visible | Magnification such that region of interest fills > 30% of screen by area | Trachea visible | Aorta and arterial duct seen in join in V shape |

Each image was reviewed and scored by two experts using a secure online platform (labelbox.com) ^33^ using a scoring guideline (outlined below). The experts scoring each image were blinded to the method used to acquire each image, and they were presented in a random order. For the clinically acceptable variable, if the two experts did not agree (i.e. one expert graded the image as clinically acceptable, and the other as clinically unacceptable), then the image was graded a third time by an experienced research sonographer using the same platform, blinded in the same way. The final score was a majority vote between the three. For the overall quality score, the mean of the scores from the two experts was used as the final score.

## Supplementary information 5: data loss

Due to software or hardware failures during the study procedures, some data were lost and therefore not available for analysis. For the paired analysis comparing the two methods, only data where a paired datapoint from each method were available were used, unpaired data were discarded. In total there were two pregnant participants who underwent only AI-assisted scans as the sonographer assigned to the manual scan was available, these have been removed from further analyses. The remaining 78 underwent scanning with both methods, and were included in the main outcome measure of diagnostic performance, and also scan duration and sonographer cognitive load. There were 11 pregnant participants in whom either the manual and/or AI-assisted reporting stage (including reporting and image saving) were affected by technical failures. The remaining 67 were included in further analyses. If a technical failure meant that a contemporaneous written report was not possible, a verbal report was recorded by the study team about any suspected malformations and the planned outcome so that the primary outcome measure could still be recorded for these scans. A data loss flow diagram is shown in Figure 8.

**Figure 8:**
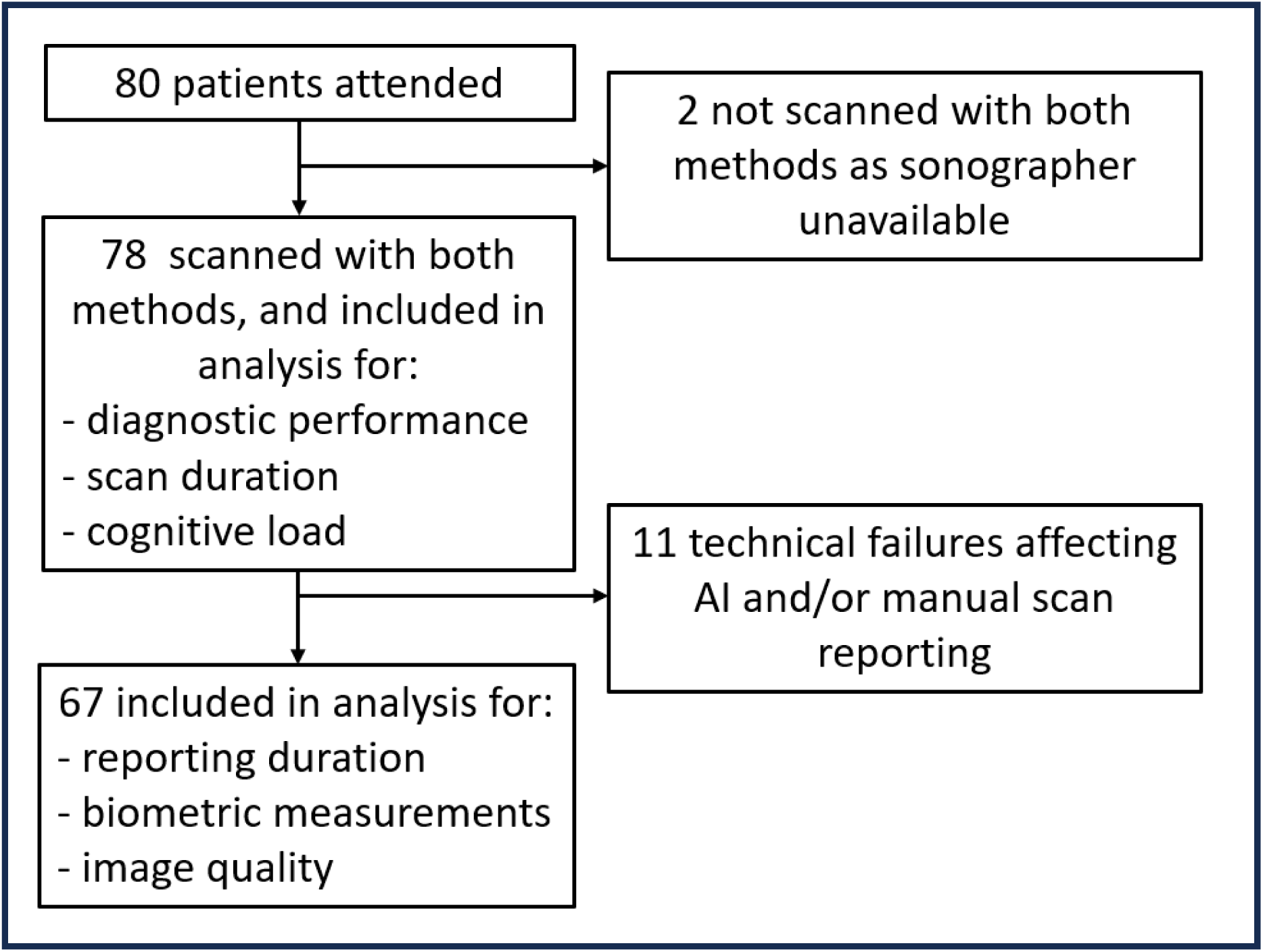
flowchart demonstrating data loss and inclusion in various analyses.

